# An Electronic Health Record-Wide Association Study to identify populations at increased risk of *E. coli* bloodstream infections

**DOI:** 10.1101/2025.05.15.25327689

**Authors:** Emma Pritchard, Karina-Doris Vihta, Samuel Lipworth, Koen B. Pouwels, Nicole Stoesser, Russell Hope, Berit Muller-Pebody, T. Phuong Quan, Jack Cregan, Colin Brown, Susan Hopkins, David W. Eyre, A. Sarah Walker

## Abstract

**Background:** *Escherichia coli* bloodstream infections (BSIs) have been under mandatory surveillance in the UK for fifteen years, but cases continue to rise. Systematic searches of all features present within electronic healthcare records (EHRs), described here as an EHR-wide association study (EHR-WAS), could potentially identify under-appreciated factors that could be targeted to reduce infections.

**Methods:** We used data from Oxfordshire, UK, and an EHR-WAS method developed for use with large-scale COVID-19 data to estimate associations between *E. coli* BSI cases, hospital-exposed controls, and 377 potential risk factors using Poisson regression models adjusted for potential confounders for three two-year financial year (FY) periods.

**Findings:** FY2022/23-2023/24 analysis included 757 (0·3%) cases and 276,758 (99·7%) controls. We identified six broad disease areas associated with increased and decreased risk of *E. coli* BSIs.

Selected renal/urological/urinary tract infection-related variables had the largest impact on risk, with 47% risk theoretically removed if prevalence of these factors could be minimised. Cancer-related variables were also associated with higher *E. coli* BSI risk (e.g. 1·20 times higher (95%CI 1·08-1·34) for every three months closer chemotherapy was delivered in the last year) as were gastrointestinal disease-related and infectious disease-related variables. Selected cardiac/respiratory-related variables were associated with lower *E. coli* BSI risk, whereas healthcare exposure had mixed effects. Associated factors varied across periods; however, broad groups were similar.

**Interpretation:** Applying an EHR-WAS approach, we show *E. coli* BSIs are largely driven by known risk factors and frailty, potentially explaining why enhanced surveillance has not reduced incidence.

**Funding:** National Institute for Health Research Health Protection Research Unit in Healthcare Associated Infections and Antimicrobial Resistance at the University of Oxford in partnership with UK Health Security Agency (NIHR200915).

## Introduction

*Escherichia coli* (*E. coli*) bloodstream infections (BSIs) have been under mandatory surveillance for fifteen years in the UK, but cases continue to rise.^1^ In contrast, enhanced surveillance of methicillin-resistant *Staphylococcus aureus* (MRSA) and *Clostridioides difficile* infection led to cases declining^2^ by providing epidemiological insights informing targeted infection prevention strategies.^3^

The UK *E. coli* BSI surveillance programme collects data on age, sex, and antibiotic resistance.^4-6^ Electronic health records (EHRs) offer a unique opportunity to identify populations at risk of infection more comprehensively, with regularly updated data streams facilitating this cheaply. Systematic searches of all features present within EHRs, described here as an EHR-wide association study (EHR-WAS, akin to genome-wide association studies) could identify novel, underappreciated factors to target to reduce infections.

Studies of infection using EHR data benefit from increased regional and national linkage between microbiology data and patient admissions driven by COVID-19,^7^ allowing more accurate laboratory-confirmed infection ascertainment and assessment of more diverse and higher numbers of associated factors, whether causal or proxies. As laboratory results, vital sign measurements, and blood test results are often automatically uploaded to electronic systems, and reasons for inpatient admissions are coded immediately after hospital discharge, EHR data should be relatively up-to-date, enabling continuous monitoring of contemporaneous factors.

Ad hoc studies have investigated *E. coli* BSI risk factors in hospital populations but often considered a limited number of factors selected a priori and were not designed for continuous monitoring.^8,9,10^ These studies found populations at highest risk included individuals on dialysis, renal disease/failure patients, cancer patients,^8,9^ and individuals with urinary catheterization/incontinence,^9^ urinary tract infections (UTI),^10^ and higher comorbidity scores.^10^

Identifying more associated factors could help target interventions by directly removing or reducing a causal mechanism or protecting high-risk groups. For example, although a Phase 3 trial of a prophylactic extraintestinal pathogenic *E. coli* vaccine has recently been halted,^11^ there is substantial ongoing research in this area. Importantly, analogous to machine learning, to target interventions, associations may not need to be causal, providing the underlying mechanisms they represent are generalizable.

Using EHRs, we aimed to identify populations at increased risk of *E. coli* BSIs using a novel EHR-WAS method which could be applied repeatedly over time, using a hospital-exposed control population to minimize missing data.

## Methods

We used the Infections in Oxfordshire Research Database (IORD): a data warehouse including inpatient admissions, outpatient appointments, emergency department (ED) visits to four large teaching hospitals serving a population of 755,000; vital signs taken during hospital attendance; microbiology (positive and negative results) and biochemistry/haematology results. The hospital group provides all acute services to the region and all community and hospital laboratory and microbiology testing. IORD has approvals from the National Research Ethics Service South Central-Oxford C Research Ethics Committee (19/SC/0403), Health Research Authority and Confidentiality Advisory Group (19/CAG/0144) as a deidentified database without individual consent.

We included all admissions from 01-April-2018 to 31-March-2024, divided into three two-year periods (April to March, keeping winter months together): FY2018/19-2019/20 (i.e., 01-April-2018 to 31-March-2020), FY2020/21–2021/22, and FY2022/23–2023/24. We looked back up to five years for potential factors, hence including data from 01-April-2013.

### Analysis cohort definition

Potential cases were all patients with *E. coli* cultured from blood (**Figure S1**). Potential controls were all individuals who were not a case and had contact in the current period (inpatient episode, outpatient or ED visits, blood test, or microbiology sample) (**Figure S2**). We included one observation per person per calendar period, selecting the first positive culture for cases and the last observation otherwise.

As many characteristics (e.g. those based on diagnosis or procedure codes) were recorded in inpatient episodes, cases and controls were included if they had an inpatient episode in the last 5y that was not attributable to the *E. coli* BSI (episode ending >72 hours (h) before blood culture collection) to minimise reverse causality as many BSIs lead to admissions and factors identified from these episodes may be consequences, not causes, of infection (20% controls had inpatient episodes ≤72h before their most recent record and were retained for analyses). Consequently, we are estimating risk of *E. coli* BSIs versus healthcare contact for other reasons.

### Identifying EHR-wide associations

We defined six variables adjusted for in all models regardless of the magnitude of association (“core” variables): age, sex, ethnicity (white vs non-white as small numbers in the latter), deprivation (Index of Multiple Deprivation (IMD) percentile),^12^ rural/urban classification, and catchment percentage (percentage of individuals in the local area visiting an Oxfordshire hospital; 0 = none, 100 = all).^13^

377 “screening” characteristics from various EHR data sources were defined from previously published research, clinical advice, and data availability (**Supplementary Methods**, definitions at https://github.com/EmmaPritchard/EHR-Risk-Factor-Definitions). Information from the 72h before blood culture collection for cases was excluded to avoid reverse causality.

Variables were included as categorical and continuous parameterisations, capturing ever/never having a characteristic (within the last 5y) and proximity of the most recent record to the current contact. Categorical effects included three levels: (i) factor recorded in the last 365 days (365d); (ii) factor recorded >365d-5y ago; (iii) factor not recorded in IORD in the previous 5ys. Continuous effects denoted days since the most recent record ≤365d ago. For inpatient admissions, outpatient appointments, ED visits, blood cultures, and urine tests, the number of occurrences (and length of stay for admissions) within the last 365d were included as variables, totalling 704 variables from 377 factors.

### Statistical Analyses

Analyses were based on previously published methodology applied to COVID-19.^14^ In brief (details in **Supplementary Methods**), for each period, starting with FY2022/23-2023/24, associations between *E. coli* BSIs (binary yes/no, cases/controls) and “core” variables were estimated using Poisson regression (log link) with cluster robust standard errors (requiring complete data for “core” variables), considering non-linearity and pairwise interactions. Each “screening” variable was then added individually to the “core” model to reduce data loss and substantial collinearity from including >700 variables. Non-linear effects of all continuous variables were considered, and levels of categorical variables grouped based on Wald tests (p<0·05). Correlation between all variables with a univariable global p<0·25 was calculated, excluding one variable from each pair with Spearman correlation coefficient>0·75 to reduce collinearity (based on clinical interpretability). We then used backwards elimination on variables with univariable global p<0·25 (to avoid missing important variables^15^) to identify a final model (exit p-value=0·05 for main effects and 0·01 for non-linear terms, keeping the linear component in the latter), grouping categorical variables during the process as above. Gross collinearity in the final model was assessed by identifying variables where the direction of effect switched between univariable and multivariable models when p<0·05. Collinear variables were kept unless the sign change was clearly influenced by another variable that conflicted with clinical or epidemiological reasoning (assessed on a case-by-case basis). The final model was refitted on complete cases for all selected variables.

The full model fitting process was repeated for FY2018/19-2019/20 and FY2020/21-2021/22, comparing selected variables across the three periods. Variables selected within each period were fit to all other periods to assess consistency of explained variation, summarising using pseudo-R-squared values.^16^

### Measuring importance

To quantify the importance of each variable and multiple variables within disease groups in multivariable models (jointly considering prevalence and predicted risk), we calculated the percentage of risk removed from the population, assuming minimal prevalence in cases and controls (**Supplementary Methods**).

### Targeting future interventions

We assessed how our models could be used to target future, hypothetically uniformly effective interventions, e.g. vaccination. We predicted the probability of having an *E. coli* BSI from the final model for each individual, identifying the optimal threshold using the Youden index (for illustrative purposes) on a receiver operating characteristic curve. We considered how using this model-derived threshold, or 10 arbitrary criteria based on age and associated factors, as criteria for vaccination would affect sensitivity, specificity, and number vaccinated (**Supplementary Methods**).

## Results

### Results from FY2022/23-2023/24

There were 953 potential cases and 276,758 controls in FY2022/23-2023/24 (**Figures S1, S2**). 196 (21%) cases were excluded due to no prior inpatient episode <5y ago (higher proportion outside hospital catchment and missing core variable data [**Table S1**]), leaving 757 cases for analysis (445 [59%] community-onset community-associated, 187 [25%] community-onset healthcare-acquired, 125 [17%] hospital-onset healthcare-associated as defined in^17^).

After fitting the core model (**Figure S3**) plus each of the 377 factors, backwards elimination on those with p<0·25 and investigating collinear variables (**Supplementary Results**), 51 variables were selected for the final multivariable model.

Selected infectious disease-related variables were associated with higher *E. coli* BSI risk (**Table 1, Figure 1**). Diagnosis codes for previous lymphadenitis or microbiology tests for CMV or EBV screening ≤1y ago were associated with higher risk: incidence rate ratio (IRR) versus >1y ago/never in EHR 1·7 (95% CI: 1·2-2·6) and 2·1 (1·3-3·2), respectively. Previous non-HIV infection diagnosis codes or blood cultures taken ≤5y ago were associated with higher risk. Skin infections or septicaemia diagnosis codes 1y ago were associated with higher risk, however, risk reduced closer to the last record (**Figure S4**). Higher and lower lymphocyte levels were associated with higher risk versus the median (**Figure S5**).

**Table 1:** Percentage of risk removed assuming minimal prevalence across all individuals (cases and controls).

| Disease area group | % risk removed assuming minimal prevalence* | Variables in disease area group | Prevalence in cases, % [n] | Prevalence in controls, % [n] |
| --- | --- | --- | --- | --- |
| **Renal/urological/UTI** | 47% | Prosthesis insertion into ureter, urine culture taken, paralysis, urine positive for *E. coli*, kidney/ureter/biliary disease, acute renal failure, urine catheter, potassium, fluid/electrolyte disease | 89% [570] | 58% [97,897] |
| **Gastrointestinal/biliary** | 36% | Albumin, bile duct/liver surgery, alcohol dependency, digestive/anal/rectal conditions, biliary tract disease, MR pancreatic region, inpatient admission under gastroenterology | 40% [258] | 22% [36,006] |
| **Infectious diseases** | 30% | Skin infection, CMV/EBV screen, lymphadenitis, sepsis, non-HIV infection, blood culture taken, lymphocytes | 73% [466] | 38% [63,661] |
| **Other** | 15% | Cataract procedure, central venous catheter, weight, haemoglobin, skin disorders | 31% [197] | 13% [20,951] |
| **Healthcare visits** | 14% | Emergency inpatient admission, inpatient admission under geriatric consultant, inpatient admission under general surgery, complex inpatient admission, emergency department visit, number of complex inpatient admissions, palliative care | 84% [535] | 55% [92,622] |
| **Cancer** | 8% | Pancreatic cancer, bone marrow transplant, extraction of bone marrow, chemotherapy, CT chest abdomen and pelvis | 19% [121] | 3% [5,240] |
| **Cardiac/respiratory** | 8% | Myocardial perfusion scan, pneumonia, upper respiratory disease, respiratory failure, COVID-19 test | 80% [512] | 59% [94,391] |

*\*calculated by taking the difference between the predicted probability of being a case given recorded exposure and the predicted probability with minimal exposure (absence of factor) in cases and controls, then dividing by the predicted probability of being a case given recorded exposure (Supplementary Methods).*

**Figure 1:**
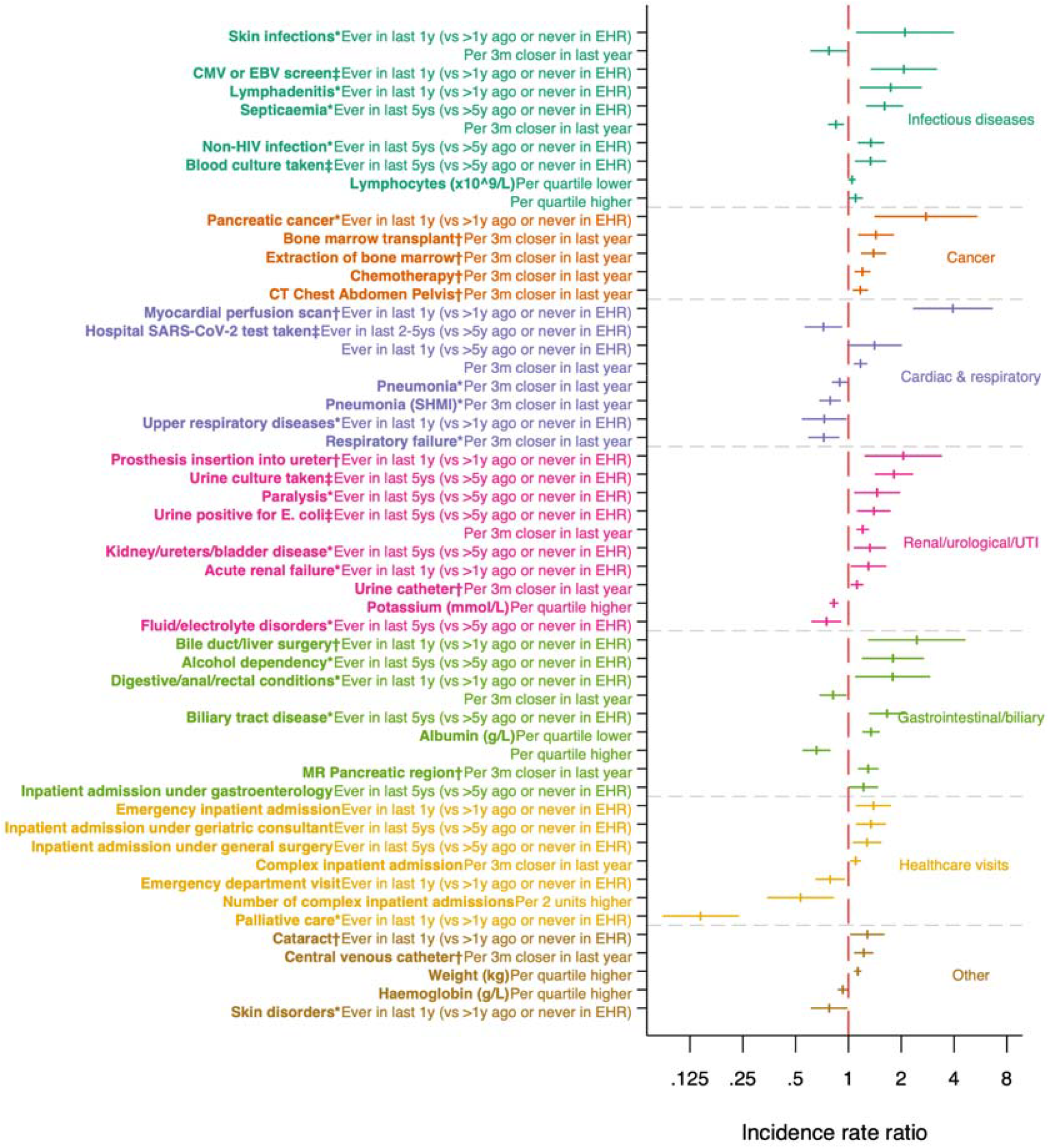
Adjusted associations (incidence rate ratios with 95% CIs) between selected characteristics and *E. coli* BSI risk in FY2022/23-2023/24. Note: factor calculated from: **\*** diagnosis codes, **†**procedure codes, **‡**microbiology data. SHMI=Summary Hospital-level Mortality Indicator. CT Chest Abdomen Pelvis is grouped with “cancer” for illustrative purposes however it may not always be cancer-related.

Several cancer-related variables were associated with higher *E. coli* BSI risk (**Figure 1, Table 1**), specifically pancreatic cancer diagnosis codes (IRR=2·8 versus >1y ago/never in EHR [95% CI 1·4-5·4]), more recent bone marrow transplant, extraction of bone marrow, chemotherapy, or computed tomography (CT) of the chest, abdomen, and pelvis.

Most selected renal/urological/UTI-related variables were associated with higher *E. coli* BSI risk (**Figure 1, Table 1**), e.g. prosthesis insertion into the ureter and acute renal failure diagnosis codes ≤1y ago versus >1y ago/never in the EHR. Urine cultures and urine positive for *E. coli* in the last 5y were associated with higher risk, with risk increasing further over the last year for the latter (from 1·4 (95% CI: 1·1-1·7) 1y ago to 3·0 (2·3-3·9) 3d ago, **Figure S4**). In contrast, fluid/electrolyte disorder diagnosis codes ≤5y ago were associated with lower risk (IRR=0·75 (0·62-0·91); defined from Summary Hospital-level Mortality Indicators, including various diagnoses, e.g. volume depletion, hyperkalaemia, and hyperosmolality). Due to high amounts of missing data for HbA1c test results (typically measured in individuals with pre-diabetes/diabetes), HbA1c was excluded from backwards elimination to reduce model instability (**Table S3**). However, as it was highly significant univariably, we added on top of the final multivariable model, with higher HbA1c associated with higher *E. coli* BSI risk (IRR 1·14 (95% CI: 1·08, 1·20) per 5mmol/mol higher), even within normal HbA1c range (**Figure S6**).

Most selected variables related to gastrointestinal disease were associated with higher *E. coli* BSI risk (**Table 1, Figure 1**), e.g. alcohol dependency or biliary tract disease diagnosis codes ≤5y ago versus >5y ago/never in EHR. Digestive/anal/rectal condition diagnosis codes ≤1y ago were associated with higher risk, but risk reduced closer to the last record. More recent pancreatic region magnetic resonance scans were associated with higher risk (IRR=1·3 (95% CI 1·1-1·5) per 3 months closer). Lower albumin was associated with a higher risk, plateauing from 36g/L onwards (**Figure S5**).

Most selected cardiac/respiratory-related variables were associated with lower *E. coli* BSI risk (**Table 1, Figure 1**), including more recent pneumonia or respiratory failure diagnosis codes in the last year. However, myocardial perfusion scans ≤1y ago were associated with a higher risk (IRR=3·9 >1y ago/never in EHR (95% CI 2·3-6·6)).

Selected healthcare exposure variables were associated with higher and lower risk of *E. coli* BSIs (**Figure 1, Table 1**). Emergency inpatient admissions, inpatient admissions under geriatric consultants, and general surgery inpatient admissions were associated with higher risk, while ED visits were associated with lower risk (after adjusting for other factors). More recent complex inpatient admissions were associated with higher risk; however, risk reduced by 47% (95% CI 17%-66%) per two additional complex inpatient admissions. Palliative care diagnosis codes were associated with lower risk (IRR=0·14 (0·09-0·24)), possibly reflecting differential ascertainment in this group.

Other variables were also associated with higher and lower *E. coli* BSI risk, e.g. more recent central venous catheter procedure codes in the last year and higher weight were associated with higher risk, skin disorder diagnosis codes (within the last year) and higher haemoglobin were associated with lower risk (**Table 1, Figure 1**).

### Risk attribution

Impact on population-level attributable risk generally increased with prevalence in cases (**Figure 2**). Renal/urological/UTI-related factors had the largest effect; 47% of the risk in cases would theoretically be removed if the whole population was like the individuals with minimal exposure (i.e. >5y ago/never) across all variables in this group (**Table 1**). These factors were highly prevalent: 89% of cases had at least one renal/urological/UTI-related factor versus 58% of controls. 87% of cases and 54% of controls had a urine culture taken in the last 5ys, with overall risk reducing by 37% if the whole population was like the individuals with no urine sample taken for culture in the last 5ys (**Figure 2**). Gastrointestinal/biliary disease-related factors had the second-largest impact, with 40% of cases having at least one associated factor in this category, followed by infectious disease-related factors. Although cancer-related variables had high IRRs, their impact on risk removal was smaller, with 8% reduction assuming the whole population was like the individuals without cancer-related variables, likely due to lower prevalence (19% cases, 3% controls).

**Figure 2:**
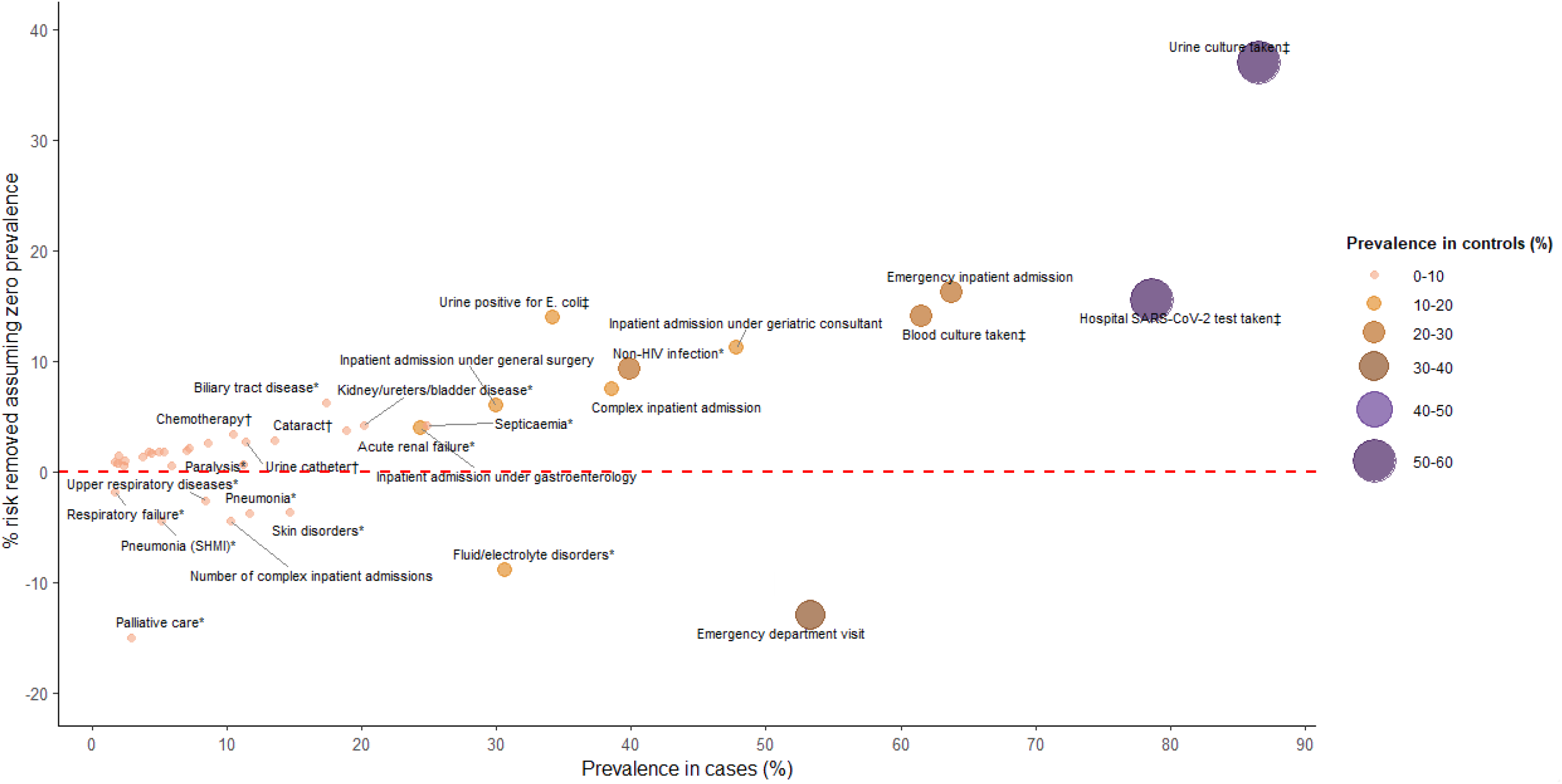
Percentage of risk removed assuming minimal prevalence across all individuals (cases and controls) vs. prevalence of each factor in case in FY2022/23-2023/24. Note: factor calculated from: **\*** diagnosis codes, **†**procedure codes, **‡**microbiology data. The red dashed line indicates no change in risk by assuming minimal prevalence in cases and controls. Blood tests and traits are not shown on the above graph as there is no corresponding prevalence. They have the following risk removal: albumin 25%, haemoglobin 13%, lymphocytes 4%, weight −0·1%, potassium −3%.

### Targeting vaccination

Using the model-derived threshold to assign a hypothetically uniformly effective, targeted intervention, such as vaccination, yielded high combined sensitivity and specificity, with 88% of controls not selected for vaccination (specificity) and 77% of cases selected for vaccination (sensitivity) (**Figure 3, Figure 4**). In contrast, simply using age-based thresholds lowered both sensitivity and specificity. 66,834 people would be selected for vaccination using an age≥65y criteria, compared with 41,676 and 20,909 using age≥75y and model-derived thresholds, respectively.

**Figure 3:**
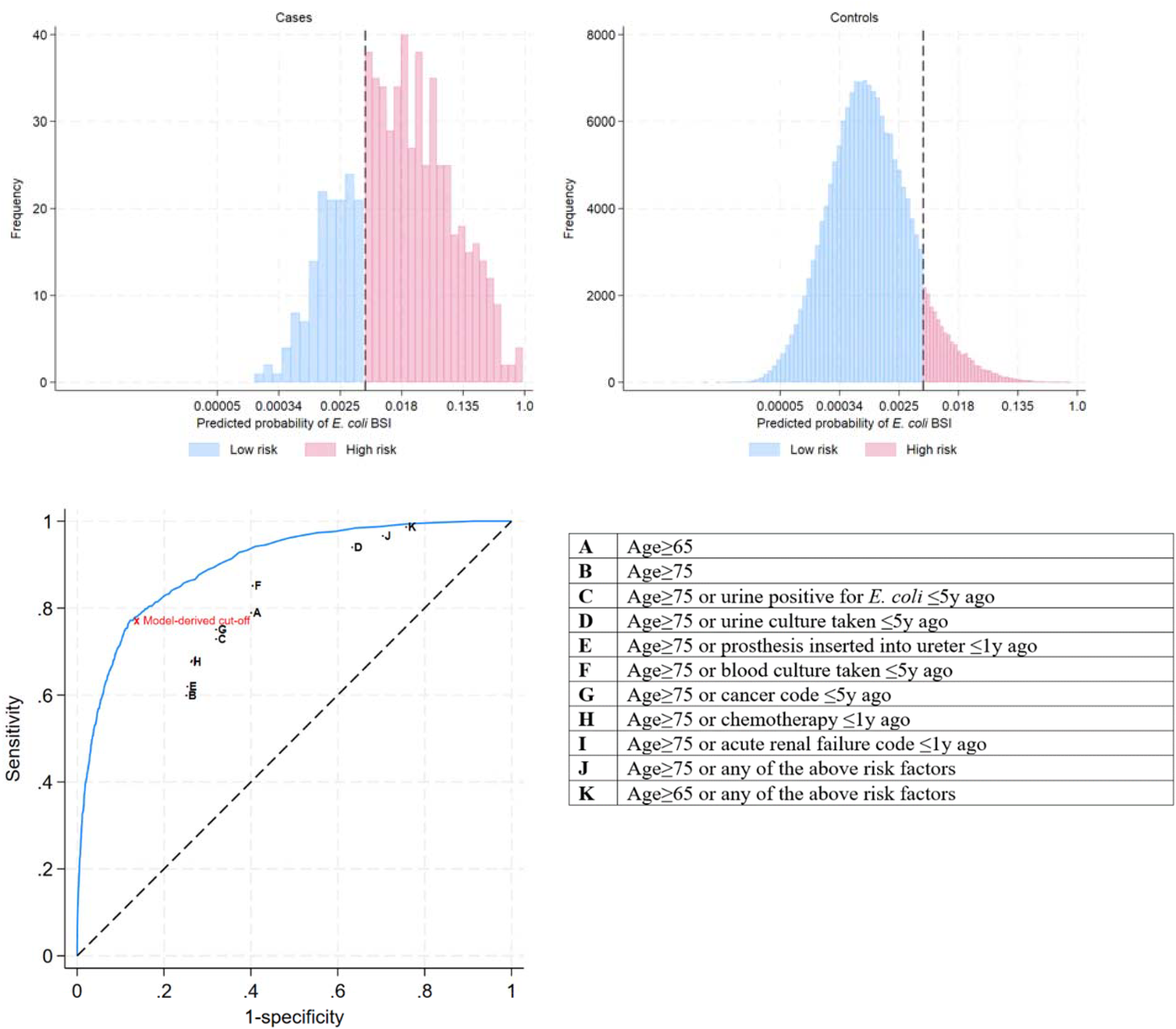
The predicted probability distribution in the low- and high-risk groups, as defined using the Youden index for cases (top left panel) and controls (top right panel), and Receiver Operating Characteristic (ROC) curve for the predicted probability from logistic regression model with vaccination criteria marked using letters (bottom panel). Note: Area Under the Curve (AUC) = 89·8% (95% CI: 88·6%-91·0%).

**Figure 4:**
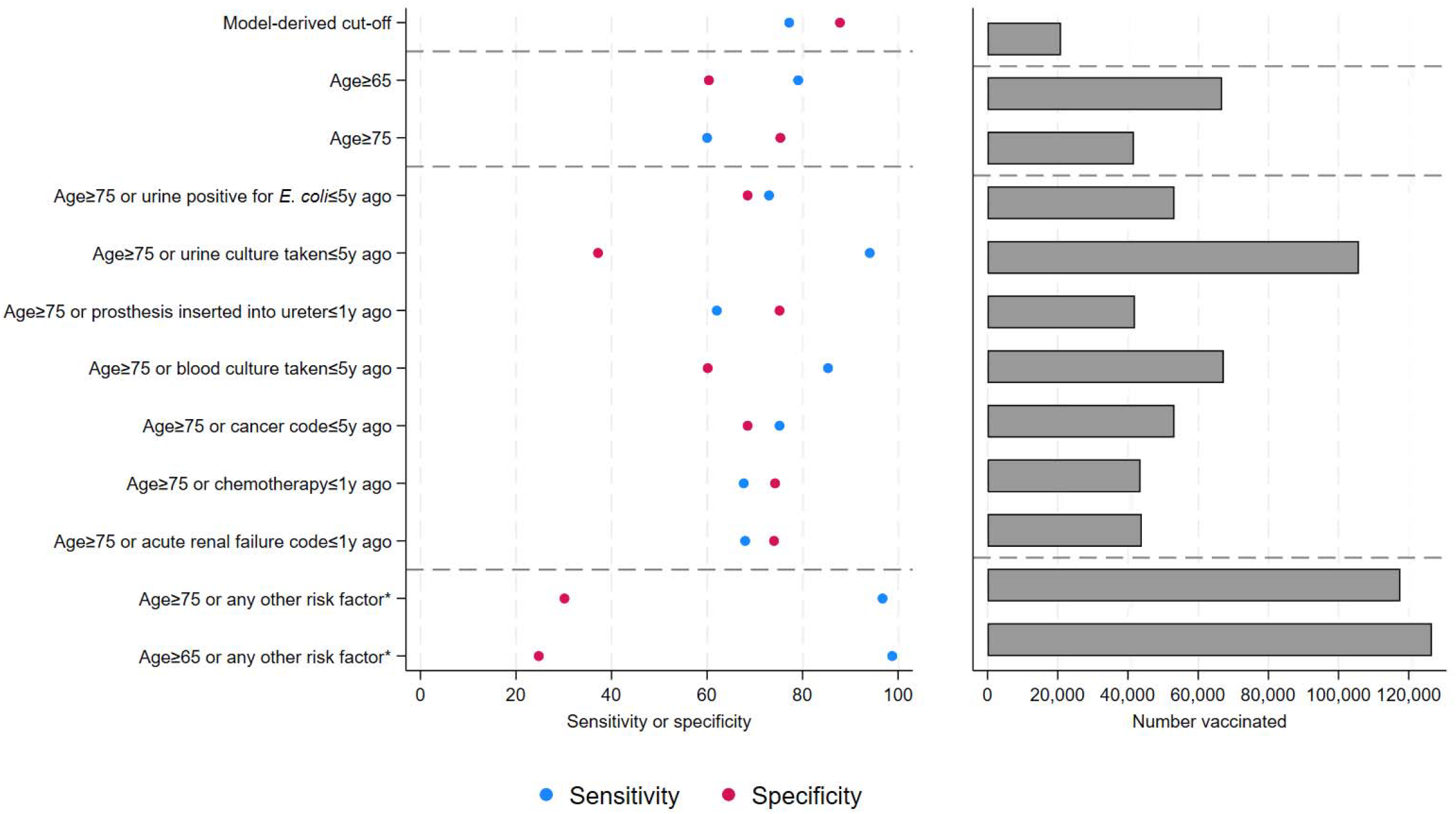
Sensitivity and specificity (left) and the number of people vaccinated with a hypothetical uniformly effective vaccine (right) using different vaccination criteria. *Any of the seven individual risk factors presented in the main panel, specifically urine positive for E. coli (≤5y ago), urine culture taken (≤5y ago), prosthesis inserted into the ureter (≤1y ago), blood culture taken (≤5y ago), diagnosis code for cancer (≤5y ago), procedure code for chemotherapy (≤1y ago), or any diagnosis code for acute renal failure (≤1y ago). Note: For clarity, horizontal dashed lines separate model-derived cut-offs, age groups, specific risk factors, and all risk factors combined.

Vaccinating those with specific associated factors, plus those aged≥75y, increased sensitivity, but lowered specificity by varying amounts, e.g. additionally vaccinating those with urine cultures taken ≤5y ago increased sensitivity to 94%, but dropped specificity to 37%. Adding in those with blood cultures taken ≤5y ago increased the sensitivity (85%) and reduced specificity by a smaller amount (60%). Vaccinating those aged≥75y or with any of the seven associated factors returned low specificity (30%) and high sensitivity (97%), vaccinating more people (n=117,564). Applying the FY2022/23-2023/24 vaccination threshold to FY2018/19-2019/20 and FY2020/21-2021/22 outperformed all other criteria (**Figure S7**).

### Results from other calendar periods

While different individual variables were identified across the three two-year periods, similar categories were consistently selected (**Figure 5**). In total, 113 variables were selected after backwards elimination in one or more periods, 83 (73%) in only one two-year period, 19 (17%) in two periods, and 11 (10%) in all three periods (**Table S5**). Variables selected in all three two-year periods had strong and high associations with *E. coli* BSIs, including urine positive for *E. coli*, blood culture taken, and chemotherapy. Many variables only selected in one period had similar characteristics selected in other periods; e.g., renal failure diagnosis codes in FY2018/19-2019/20 versus acute renal failure diagnosis codes in FY2020/21-2021/22 and FY2022/23-2023/24, and any cancer or rectal cancer diagnosis codes in FY2020/21-2021/22, liver cancer or rectal cancer diagnosis codes in FY2018/19-2019/20, and pancreatic cancer diagnosis codes in FY2022/23-2023/24. The largest difference was the increase in the cardiac/respiratory groups from FY2020 due to COVID-19, resulting in large reductions in risk removed in FY2020/21-2021/22, likely due to changes in hospital population composition (**Figure 5**). Further, factors related to neurological/psychosis disorders were identified in FY2018/19-2019/20 and FY2020/21-2021/22 but not in FY2022/23-2023/24; however, their impact was small (risk removed=-9% and 8%, respectively) (**Supplementary Results**).

**Figure 5:**
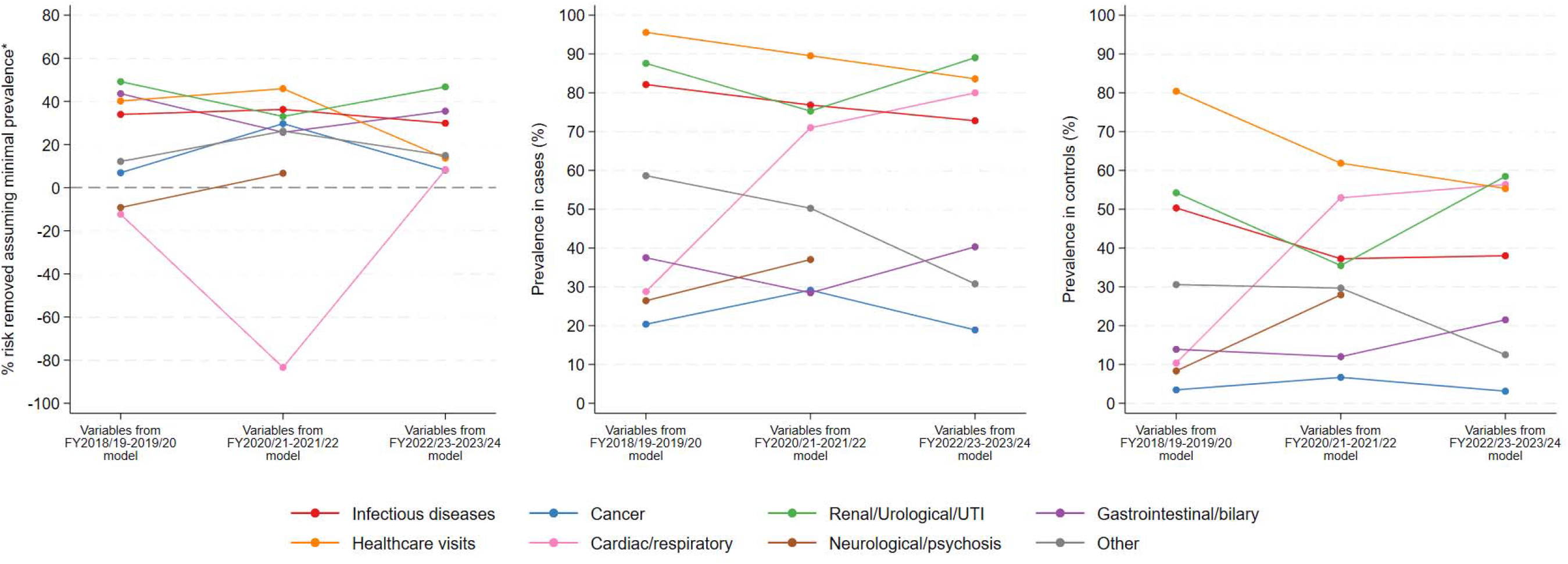
Percentage of risk removed assuming minimal prevalence across all individuals (cases and controls) (left), and prevalence in cases (middle) and controls (right) across FY2018/19-2019/20, FY2020/21-2021/22, and FY2022/23-2023/24. *calculated by taking the difference between the predicted probability of being a case given recorded exposure and the predicted probability with minimal exposure (in cases and controls), then dividing by the predicted probability of being a case given recorded exposure (**Supplementary Methods**). Negative means risk increased at the population level.

Prevalence of variables remained mostly stable across all periods (**Figures S14-S16**). Most variables which varied between periods related to COVID-19, e.g., increases in previous hospital SARS-CoV-2 tests, non-HIV infection diagnosis codes, and respiratory failure diagnosis codes from FY2018/19-2019/20 to FY2020/21-2021/22.

Variation explained reduced modestly fitting variables selected in FY2018/19-2019/20 (pseudo-R-squared=22.1%) to data from FY2020/21-2021/22 (pseudo-R-squared=18.2%) and FY2022/23-2023/24 (pseudo-R-squared=19.5%) (**Table S6**). Results were similar in other periods.

## Discussion

Using our EHR-WAS approach, we identified EHR-derived factors associated with *E. coli* BSIs over six years. Many associations identified reflect known risk factors, e.g., in FY2022/23-2023/24, previous infectious diseases, cancer, renal/urological/UTI, and gastrointestinal/biliary-related factors were generally associated with higher risk. Respiratory illnesses were associated with lower risk of *E. coli* BSI, likely reflecting common reasons for attending hospital unrelated to *E. coli* BSIs, given controls were hospital-exposed to reduce missing data.^18^ Previous healthcare attendances were associated with higher BSI risk, e.g. previous emergency inpatient admissions, and lower risk, e.g. higher numbers of complex inpatient admissions, potentially reflecting survivor bias. *E. coli* BSI risk was associated with blood test results, including higher risk for individuals with lower albumin. Considering prevalence and predicted risk, targeting individuals with common associated factors such as previous urine cultures taken may be useful, although these were also common in controls. Associations differed across successive two-year periods, possibly due to the influence of COVID-19 on hospitalisation; however, similar groups of factors were identified, suggesting underlying risk likely remained similar.

Many factors identified in this study associated with higher *E. coli* BSI risk were common markers or contributors to frailty, e.g., low albumin, low haemoglobin, frequent previous healthcare attendances, and chronic illnesses, including renal failure, cancer, and gastrointestinal conditions. This characterization differs from MRSA and *C. difficile*, where surveillance reduced incidence, likely as these infections are caused by healthcare-associated acquisition and antimicrobial use that could be targeted by interventions. In contrast, *E. coli* BSIs often have an intrinsic origin from gut flora, and events leading to BSI are more common in those experiencing frailty. This highlights the complexity of reducing *E. coli* BSIs, with interventions having to incorporate the multifaceted nature of frailty rather than there being a “silver bullet”. Some specific factors identified could be targeted further, e.g. urinary catheters, identified here and in other studies.^9,19,20^ Avoiding unnecessary catheter use, removing catheters when no longer needed,^21^ and prioritising bladder outflow obstruction surgery could reduce risks. Patients with cancer were at increased risk,^22^ reflecting risks from surgery and chemotherapy, which form a necessary part of treatment, but where there are still opportunities to mitigate risks, e.g., implementing better hand hygiene and using full-barrier precautions during central line catheter insertion, which has previously been shown to reduce infection risk.^23^ We found higher *E. coli* BSI risk in individuals within the normal and pre-diabetic HbA1c range, suggesting that checking HbA1c levels in people with urinary infections could help identify those at increased *E. coli* BSI risk.

Using factors identified in the final multivariable models could potentially improve targeting of future prophylactic vaccines if these are effective. Compared to age-based thresholds, model-based targeting improved specificity, meaning fewer low-risk individuals would be unnecessarily vaccinated, reducing costs and any vaccine-associated risks. We assumed the vaccine would be uniformly effective; however, it may be less effective in highest-risk groups with impaired immune responses, reducing the number of BSIs prevented (**Figure S17**). We used the Youden index to identify a model-based cut-off, balancing sensitivity and specificity, which may not be ideal. While Decision Curve Analysis^24^ could be more appropriate, determining how to balance costs and benefits of this hypothetical vaccination is unclear.

While our models estimated associations between factors and *E. coli* BSIs rather than causal relationships, these associations could still inform interventions, provided they are reliable and generalizable. Targeting interventions at broader groups with higher overall risk rather than focusing on specific procedures may be effective. For example, we found recent pancreatic region magnetic resonance scans were associated with higher BSI risk. While this does not imply causation, these individuals may have some underlying characteristic putting them at higher risk, therefore, interventions could be targeted at these patients. This approach highlights the potential for using associations to guide public health strategies, even without direct causal evidence.

A challenge with our approach is that multiple features identified in EHR may represent a unified clinical pathway or condition, e.g. bone marrow aspirate, chemotherapy, bone marrow transplant, and testing for EBV/CMV may all be experienced by patients undergoing bone marrow transplants. Risks may also be captured in different ways, e.g. procedure codes for pre-operative imaging, procedure codes for surgery, and cancer diagnostic codes. This can make models more difficult to interpret and mean that different variables representing similar underlying factors were identified across different periods, as we observed. Clustering of related variables before or during model fitting could improve this.

A key study limitation was its conduct within a single hospital group, albeit large and covering around 1% of the UK population. Additionally, to reduce missing data, we restricted our analyses to individuals with inpatient episodes, excluding many individuals from the control group who were likely less comorbid, potentially causing underestimating associations. However, most cases were retained, and we observed little difference in demographic model estimates when considering a broader, healthcare-based control group.^18^ We did not have access to community prescriptions and did not include hospital prescriptions as potential factors. Previous antibiotic use has been associated with higher *E. coli* BSI incidence.^25^ Other drugs, including anti-depressants, can also affect the gut microbiome,^26^ though their impact on BSI risk is unclear. Including prescriptions may be useful if community prescribing data is available.

Another limitation was the relatively small number of cases, although we found highly significant effects. The observed variation in associated factors over two-year periods may reflect true changes or model instability, which could be assessed using bootstrapping.^27^ To increase statistical power, future studies could use larger, national-level datasets which would also improve generalizability, allow analysis stratified by subgroups of interest (e.g. community-acquired, hospital-onset cases), and reduce the risk of missing BSIs; using Oxfordshire data alone may miss individuals receiving care outside the region. Although *E. coli* BSIs are captured by UKHSA’s Second Generation Surveillance System, which has previously been linked to Hospital Episode Statistics, the national dataset would not include key factors identified in this study, such as blood tests, vital signs, and culture-negative microbiology results. Selecting cases and controls while balancing missing data and bias is complex. We excluded cases without previous inpatient episodes; their lower hospital interactions may mean different risk factors and interventions are needed. However, without access to primary care data, many factors for these individuals could not be derived. Future studies could investigate factors in populations with varying amounts of healthcare contact.

Overall, we found that *E. coli* BSIs were largely associated with known risk factors and frailty, explaining why enhanced surveillance has not led to reduced incidence. Our study also demonstrates an EHR-WAS approach that can be used with EHR data to identify associated factors without constraining the search by prior knowledge. This may have applications to other infectious diseases and particularly how associated factors change over time.

## Research in Context

### Evidence before this study

Electronic health records (EHRs) offer an opportunity to exploit rich data on potential risk factors linked to microbiological isolations which could be used to target inventions to reduce *Escherichia coli* (*E. coli*) bloodstream infections (BSIs). We searched PubMed up to 14th February 2025 for epidemiological journal articles using the terms (“Escherichia coli” OR “E. coli”) AND (“bloodstream infection*” OR “bacteremia” OR “septicemia” OR “bacteraemia” OR “septicaemia”) AND (“electronic health record*” OR “EHR” OR “electronic medical record*” OR “EMR”) AND (“risk factor*” OR predictor*) without data or language restrictions. Most studies investigated a limited number of risk factors defined a priori and were not designed for continuous monitoring. Further, many were conducted on a specific population of interest, for example, acute leukaemia patients or patients receiving haemodialysis. Urinary catheterisation, higher co-morbidities, and dialysis were commonly reported risk factors in these studies; however, none considered a large number of factors agnostically to identify new associations and reduce bias.

### Added value of this study

In a large EHR dataset including microbiological isolations linked to hospital attendances, we identified factors associated with *E. coli* BSIs from a pool of over 700 agnostically defined variables. We identified 51 associated factors in the most recent two years including factors previously identified (e.g. previous *E. coli* positive urine) and new factors generally related to frailty (e.g. albumin). Using a novel method previously developed for COVID-19 data, we were able to compare associated factors in the previous two non-overlapping two-year periods, showing how, while individual associated factors varied, broad groups of factors remained similar. We also demonstrated how carefully built multivariable models could be used to target potential future interventions such as vaccines, balancing both sensitivity and specificity, ensuring good vaccine coverage in high-risk groups while avoiding vaccination in low-risk individuals.

### Implications of all the available evidence

This study demonstrates the potential of using large-scale routine EHRs to identify populations at higher risk of *E. coli* BSIs. By using a more agnostic approach, while confirming already known risk factors, the methodology used in this study allowed new associated factors to be identified and also demonstrated how it could be used on a continuous basis to assess changes in them. We explored how interventions could be prioritised by combining prevalence and effect estimates for more impactful interventions, and explored how vaccination could be targeted in the future using many associated factors.

## Data Availability

The datasets analysed during the current study are not publicly available as they contain personal data but are available from the Infections in Oxfordshire Research Database (https://oxfordbrc.nihr.ac.uk/research-themes-overview/antimicrobial-resistance-and-modernising-microbiology/infections-in-oxfordshire-research-database-iord/), subject to an application and research proposal meeting the ethical and governance requirements of the Database. For further details on how to apply for access to the data and for a research proposal template please.

## Author Contributions

The study was designed and planned by EP, DWE, and ASW. EP conducted the statistical analysis of the data. EP, DWE, and ASW drafted the manuscript and all authors contributed to interpretation of the data and results and revised the manuscript. All authors approved the final version of the manuscript.

### Funding

This study was funded by the National Institute for Health Research (NIHR) Health Protection Research Unit in Healthcare Associated Infections and Antimicrobial Resistance at Oxford University in partnership with the UK Health Security Agency (UKHSA) (NIHR200915) and the NIHR Biomedical Research Centre, Oxford. DWE is supported by a Robertson Fellowship. The views expressed in this publication are those of the authors and not necessarily those of the NHS, the National Institute for Health Research, the Department of Health or the UKHSA.

## Acknowledgements

This work uses data provided by patients and collected by the UK’s National Health Service as part of their care and support. We thank all the people of Oxfordshire who contribute to the Infections in Oxfordshire Research Database. Research Database Team: L Butcher, H Boseley, C Crichton, DW Crook, D Eyre, O Freeman, J Gearing (community), R Harrington, K Jeffery, M Landray, A Pal, TEA Peto, TP Quan, J Robinson (community), J Sellors, B Shine, AS Walker, D Waller. Patient and Public Panel: G Blower, C Mancey, P McLoughlin, B Nichols.

