## Supplementary Material for "An Electronic Health Record-Wide Association Study to identify populations at increased risk of *E. coli* bloodstream infections"

### 1 Supplementary Methods

#### 1.1 Defining potential risk factors

Details on how variables were selected are outlined below, with variables broadly being selected based on risk factors defined in previously published research (e.g.^3,4^), clinical advice and knowledge, and the availability of data. The full list of potential risk factors considered including definitions and links to where the definitions were previously published, if applicable, is available at <https://github.com/EmmaPritchard/EHR-Risk-Factor-Definitions>.

We used ICD-10 codes from both inpatient admissions and outpatient appointments and used various published information to define potential risk factors. First, we calculated the Charlson co-morbidity index^5^ using predefined ICD-10 codes^6^ with updated weightings from Quan et al.^7^. We considered the composite score as a continuous variable, but also the presence of the individual components separately as binary variables. We defined frailty using a pre-defined frailty score for use with EHRs by Gilbert et al.^8^ 143 potential risk factors were defined using the pre-defined Summary Hospital-level Mortality indicator groups.^9^ Potential risk factors used in previous studies including renal dialysis and palliative care were defined from previous studies looking at *E. coli* BSIs.^4^

For procedure codes (coded via OPCS-4 codes), we defined common procedures including scans such as computed tomography (CT) scans of body regions, endoscopies, and catheter use. We used published coding lists to define additional variables from procedure codes for chemotherapy,^10^ dialysis,^11^ and transplant.^12^ Eighteen specific surgery groupings (e.g. including large bowel surgery and hip replacement) were defined from previously published UKHSA guidance.^13^ This document also flagged whether each surgery within each grouping was mostly clean, clean-contaminated, or contaminated, and hence three additional groupings of surgeries under these headings were considered as potential risk factors.

We created variables from microbiological samples considering both whether a test was requested and the test result. Variables were created for pathogens commonly isolated from urine samples: urine positive for *E. coli*, Enterococcus, Klebsiella, and Enterobacterales. Similarly, for blood samples, any blood culture positive for *S. aureus* was considered as a binary variable with all other isolated pathogens having very small numbers in the *E. coli* cases and therefore not included. Any urine sample collected for culture or any blood test done were also created as variables alongside the number of both these tests requested irrespective of results as, based on previous research, requesting a test can be as predictive of ill-health as the positive test/test result itself.^14^ We also considered any of the following tests requested as variables: faeces culture (and separately those positive for *Clostridioides difficile*), COVID-19 swabs (and those positive) and influenza/Respiratory Syncytial Virus (RSV) PCRs (polymerase chain reaction tests), respiratory samples (and those positive for *Pseudomonas aeruginosa*), surface swab culture taken, screen for MRSA (Methicillin-resistant Staphylococcus aureus), screen for CMV (Cytomegalovirus) or EBV (Epstein–Barr virus), or CNS (central nervous system) culture taken.

We selected blood test results from twenty tests based on the tests more commonly done and advice from clinicians to capture a variety of clinical information. We used routine tests such as those done as part of a full blood count, including white cell count, platelets, and haemoglobin levels, to assess general health indicators.^15^ We also included tests which were markers for risk factors of interest, including CRP as a marker for inflammation, bilirubin for liver function, and urea for kidney function. Other tests were selected as they are used widely to screen for specific illnesses, including HbA1c measurements for diabetes/pre-diabetes and the Prostate-Specific Antigen test (PSA) for prostate cancer (men only). We included variables for six key vital signs: heart rate, respiratory rate, systolic blood pressure (SBP), diastolic blood pressure (DBP), oxygen saturation, and temperature. These were all tests taken routinely.

For all vital signs and blood test results, extreme values incompatible with life (based on clinician judgment) were dropped from the analysis. To avoid undue influence of extreme values, we truncated all continuous variables at the 5th and 95th percentiles for variables that did not include a natural zero (BMI, height, systolic and diastolic blood pressure, heart rate, respiratory rate, temperature, Charlson score, albumin, creatinine, haemoglobin), and at the 0th and 95th percentiles otherwise (eosinophils, neutrophils). We truncated oxygen saturation at the 5th and 100th percentiles since an oxygen saturation of 100% is within the normal physiological range.

All blood test results and vital signs were matched to their closest inpatient admission, outpatient appointment, and A&E attendance. If measurements did not fall strictly within any of these, measurements were matched to admissions/attendances within ±72 hours of their collection. A small proportion of vital signs (<1%) were not strictly within, or within ±72 hours, of a recorded inpatient admission, outpatient appointment, or A&E visit. These measurements were subsequently matched to their closest admission. A larger proportion of blood test results did not fall strictly within, or within ±72 hours of, a hospital attendance but these were expected as blood tests outside of the hospital setting (i.e. those from samples taken at GPs) were tested within the hospital and hence recorded in the EHR. These measurements were therefore left unmatched to healthcare attendances. We prioritised the closest measurements to the last contact date taken outside hospital attendances, followed by those taken during outpatient appointments, then ED visits, and inpatient admissions, arguing that these would most plausibly reflect non-acute physiological “average” values for each individual.

#### 1.2 Non-linearity and interactions

Restricted cubic splines with between one to five internal knots were considered for all continuous core variables, with a minimum of one internal knot included for age due to expected variation and linear effects allowed for deprivation and catchment percentage. Internal knots were placed at even percentiles throughout the range of values for each variable (after truncation at the 5th and 95th percentiles) with boundary knots included at the 10th and 90th percentiles of the range of values for each variable. Non-linearity was tested in univariate Poisson models for each continuous characteristic, with a Bayesian Information Criterion (BIC) difference greater than 10 resulting in non-linear parameterisation with more knots being selected.^16^

Pairwise interactions between all core variables were tested, using backwards elimination on all interactions which individually had global heterogeneity p-value<0.002 (Bonferroni adjustment, 0.05/21 (number of interaction tests)).

#### 1.3 Grouping levels of categorical variables

We used Wald tests to test whether there was evidence that the effect of having the factor ≤365d ago differed from having the factor >365d ago, combining these categories if p>0·05. Similarly, whether there was evidence that the effect of having the factor between >365d-5y ago differed from having the factor >5y ago or never in the EHR, combining these categories if p>0·05.

#### 1.4 Measure of importance

To calculate the percentage of risk removed from the population under the assumption of minimal prevalence, we used the following formula^17^:

$$\frac{\Pr\left( E. coli BSI \right)-\Pr\left( E. coli BSI \right|minimal exposure)}{Pr(E. coli BSI)}$$

The predicted probability of having an *E. coli* BSI was predicted from the final selected multivariate model, and the predicted probability of having an *E. coli* BSI under the assumption of minimal prevalence was calculated from the final selected multivariate model after setting individual variables to their minimal prevalence level. The probabilities were summed over all cases and controls, and then the relative reduction calculated.

For laboratory test variables, values were set to the mid-point of laboratory-reported reference ranges under the assumption of minimal prevalence in cases and controls.

#### 1.5 Targeting vaccination

We refitted the final model using logistic regression (ensuring probabilities were bounded between zero and one).

The following criteria were defined based on age and associated factors identified after backwards elimination: two age thresholds (age≥65y or age≥75y), and seven risk-based criteria requiring age≥75y or one of the following: urine positive for *E. coli* (≤5y ago), urine culture taken (≤5y ago), prosthesis inserted into the ureter (≤1y ago), blood culture taken (≤5y ago), diagnosis code for cancer (≤5y ago), procedure code for chemotherapy (≤1y ago), or any diagnosis code for acute renal failure (≤1y ago). A final criterion required either age≥75y or any one of the seven risk-based criteria.

### 2 Supplementary Results

#### 2.1 Results from the core model

Core variables differed between cases and controls (**Table S1).** Cases were generally older than controls. There were more women in controls (56%) than cases (48%). A higher proportion of controls were missing ethnicity (23%) compared to cases (13%). Deprivation and rural/urban classification had a similar distribution between cases and controls. The distribution of catchment percentages was right skewed for cases and controls but with controls including more individuals with lower catchment percentages on average.

In multivariable models, being of non-white ethnicity was associated with a higher risk of *E. coli* BSIs when compared with white ethnicities, although this association was not statistically significant at the p<0.05 threshold and had large confidence intervals (**Figure S3**). Being female was associated with a lower risk of *E. coli* BSIs compared with males. Residing in more deprived areas was associated with higher risk. Older age was strongly associated with an increased risk, particularly in areas with higher catchment percentages (p-value for interaction = 0·0009).

#### 2.2 Collinearity

Fifty-three variables were selected after backwards elimination (exit p>0·05). There was evidence of collinearity for some variables as measured by IRRs swapping signs between univariate (“core adjusted”) and multivariate (fully adjusted) analyses (**Table S2**). Some variables with evidence of collinearity using this metric may instead be examples of “competing risks” that reflect other reasons why individuals may be admitted to hospital and hence be in the control group. For example, fluid/electrolyte disorders may represent a common reason to come into hospital that is not associated with having an *E. coli* BSI. After adjusting for other associated factors, risk estimates are therefore negative. The effects of these variables remained broadly unchanged when other variables were removed one by one from the multivariate model, so they were retained. Other collinear variables potentially related to “healthy survivor” bias, for example, the number of complex inpatient admissions, with individuals surviving multiple complex inpatient admissions having lower risk. A diagnosis code for COVID-19 was highly correlated (0·71, but below the 0·75 threshold to remove collinear variables before model fitting) with a SARS-CoV-2 positive test result from microbiology, both of which were selected in the final model. To assess the influence of including both these variables on the final model estimates, four additional models were summarised, removing each variable from the final model, creating a combined variable representing either/or both COVID-19-related variables, and removing both variables (**Table S4**). Removing both variables returned the lowest BIC and hence both variables were removed from the final model. All other collinear variables were kept in the final model.

#### 2.3 Results from other periods

In FY2020/21-2021/22, infectious diseases, cancer, renal/urological/UTI, and healthcare visit variables were mostly associated with a higher risk of *E. coli* BSIs (**Figure S8-Figure S10**). Having a SARS-CoV-2 test taken, or a positive result, were both associated with a lower risk of *E. coli* BSIs, likely due to small numbers having SARS-CoV-2 tests before the 1st April 2020 when the period began. Cardiac/respiratory factors were associated with both increased and decreased risk. Previous inpatient admissions were associated with an increased risk of *E. coli* BSI, specifically for emergency inpatient admissions, admissions under a geriatric consultant, admissions in general surgery, and admissions lasting >8 hours.

There were some differences between the FY2020/21-2021/22 and the FY2022/23-2023/24 cohorts. First, a diagnosis code for pneumonia in the last year was associated with an increased risk of *E. coli* BSI (incidence rate ratio (IRR)=2·1 [95% 1·2-3·4]); however, this risk reduced the closer the record was to the current contact (IRR=0·52 (95% 0·35-0·77) three days before current contact, **Figure S10**). Second, more variables related to neurological or psychiatric conditions were selected in FY2020/21-2021/22. For example, having a diagnosis code for a non-organic mental disorder was associated with a higher risk of *E. coli* BSIs in FY2020/21-2021/22: IRR=1·3 (95% CI 1·0-1·5).

Again, similar characteristics were observed in the FY2018/19-2019/20 cohort but with different variables selected (**Figure S11**-**Figure S13**). Variables such as chemotherapy, previous blood cultures, and previous urine cultures were consistently selected and associated with a higher risk of *E. coli* BSIs. Differences between FY2018/19-2019/20 and other periods included a diagnosis code for diabetes being associated with an increased risk of *E. coli* BSIs: IRR=1·6 (95% 1·3-1·9).

### 3 Supplementary Figures

Figure S1:Flowchart of the case population.

FY2018/19-2019/20: **772** records, **772** individuals

FY2020/21–2021/22: **677** records, **677** individuals

FY2022/23–2023/24: **757** records, **757** individuals

Individuals excluded: no inpatient episode in the last 5 years, ending at least 72 hours before current contact:

FY2018/19-2019/20: **193** individuals

FY2020/21-2021/22: **176** individuals

FY2022/23-2023/24: **196** individuals

**3,627** *E. coli* isolations from blood between 1st April 2018-31st March 2024 from **2,775** individuals.

**2,932** *E. coli* bloodstream infections

**2,775** individuals

**695** records excluded: within 90 days of an index *E. coli* blood culture (90-day deduplication)

**69** BSIs excluded from **69** people: patients aged <16 years old at *E. coli* collection date

**0** records excluded from **0** people: missing age

**0** records excluded from **0** people: missing sex

**2,863** *E. coli* bloodstream infections

**2,706** individuals

FY2018/19-2019/20: **965** records, **965** individuals

FY2020/21–2021/22: **853** records, **853** individuals

FY2022/23–2023/24: **953** records, **953** individuals

**92** records excluded: not the first *E. coli* bloodstream infection in the two-year period

*Note: “current contact” is defined as the first positive E. coli blood culture in the period*

Figure S2: Flowchart of the potential control group population.

FY2018/19-2019/20: **276,394** records, **276,394** individuals

FY2020/21–2021/22: **260,977** records, **260,977** individuals

FY2022/23–2023/24: **276,758** records, **276,758** individuals

Individuals excluded: no inpatient episode in the last 5 years, including current contact:

FY2018/19-2019/20: **273,436** individuals

FY2020/21–2021/22: **271,525** individuals

FY2022/23–2023/24: **300,649** individuals

Inpatient contact between 1st April 2018-31st March 2024;

**N = 1,193,365,**

**Unique people = 464,985**

Outpatient appointments between 1st April 2018-31st March 2024;

**N =7,375,006**

**Unique people =803,224**

Emergency department visits between 1st April 2018-31st March 2024;

**N = 874,074,**

**Unique people =432,500**

Microbiology sample taken between 1st April 2018-31st March 2024;

**N =2,577,389,**

**Unique people =507,862**

Blood test result recorded between 1st April 2018-31st March 2024;

**N =7,609,711,**

**Unique people =701,297**

**19,629,545** records of healthcare contact between 1st April 2018-31st March 2024 from **1,039,643** individuals.

**17,672,232** records

**862,097** individuals

**1,699,563** records excluded from **196,664** people: patients aged <16 years old at healthcare contact

**108** records excluded from **86** people: missing age

**280** records excluded from **73** people: missing sex

**257,362** records excluded from **3,722** people: hospital contact after first *E. coli* BSI recorded since 1st April 2013

FY2018/19-2019/20: **549,830** records, **549,830** individuals

FY2020/21–2021/22: **532,502** records, **532,502**individuals

FY2022/23–2023/24: **577,407** records, **577,407** individuals

**16,012,493** records excluded: not the last healthcare record in the two-year period

Figure S3: Estimates from the core model for main effects (left) and the interaction between age (continuous, non-linear) and catchment percentage (continuous, linear).

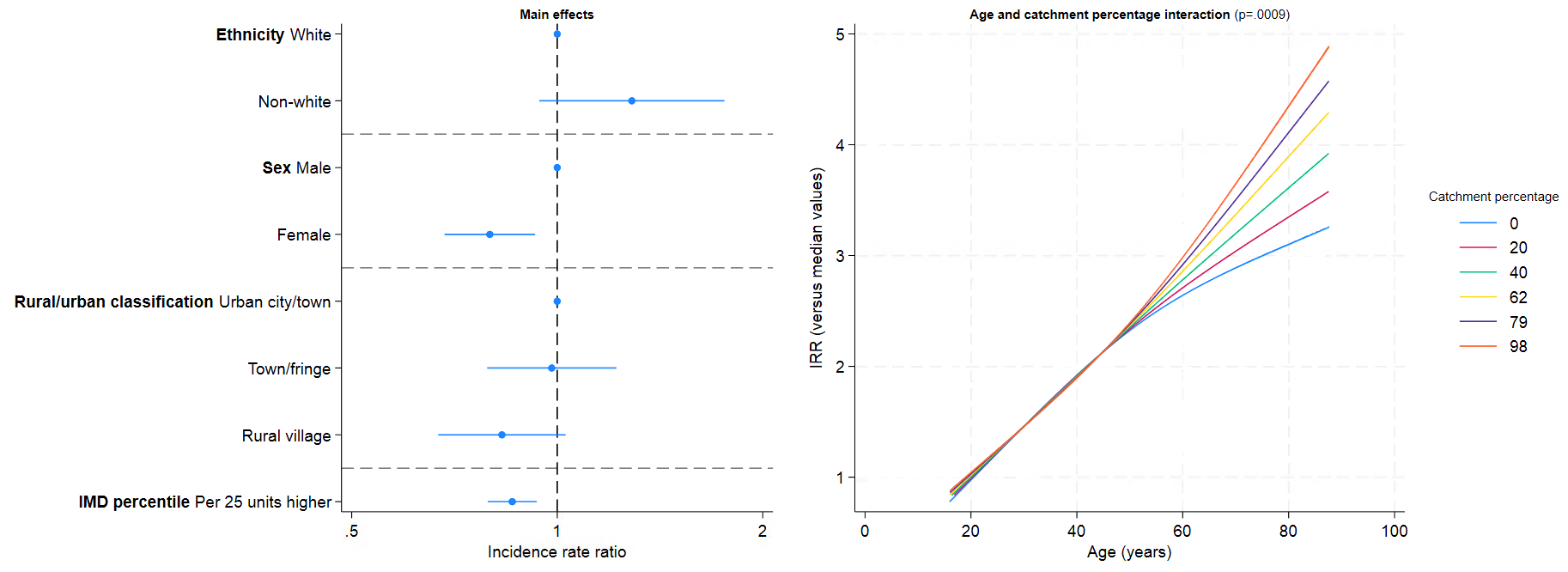

Figure S4: Associations (incidence rate ratios with 95% CIs) between characteristics where both a categorical variable and continuous linear variable were included and *E. coli* BSI FY2022/3-2023/4.

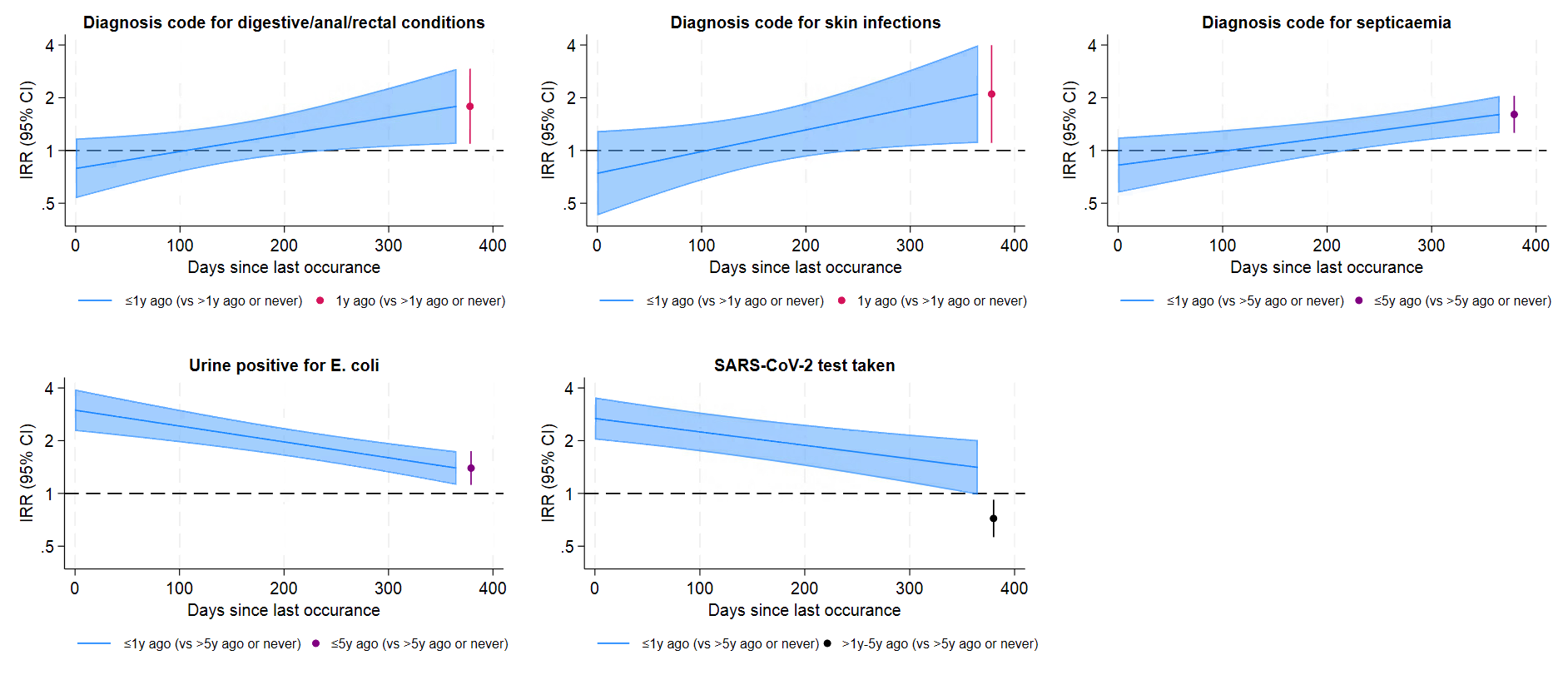

Figure S5: Adjusted associations (incidence rate ratios with 95% CIs) between continuous characteristics with non-linear effects on *E. coli* BSI in FY2022/23-2023/24.

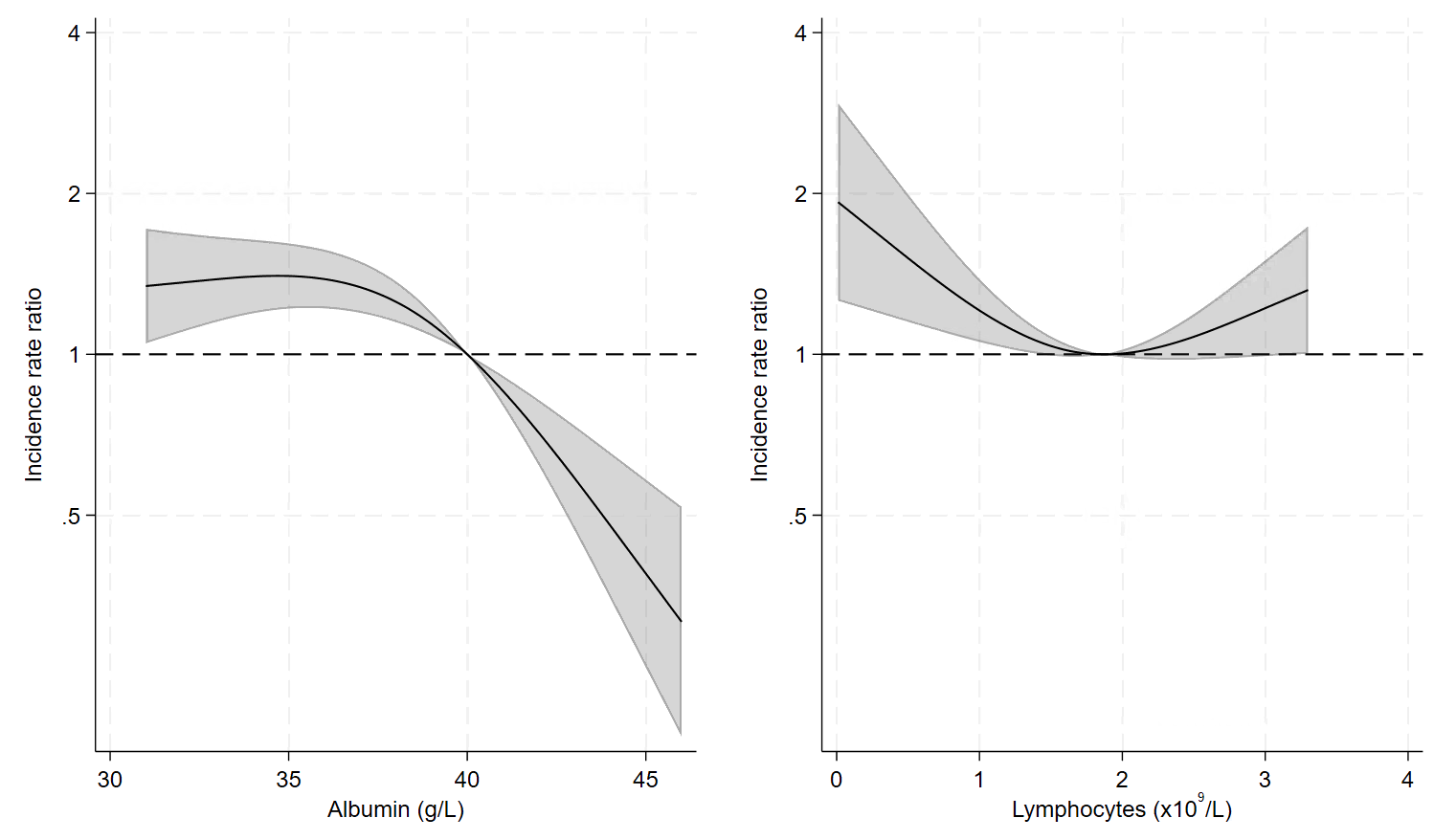

Figure S6: Adjusted association (incidence rate ratios with 95% CIs) between HbA1c and *E. coli* BSI risk.

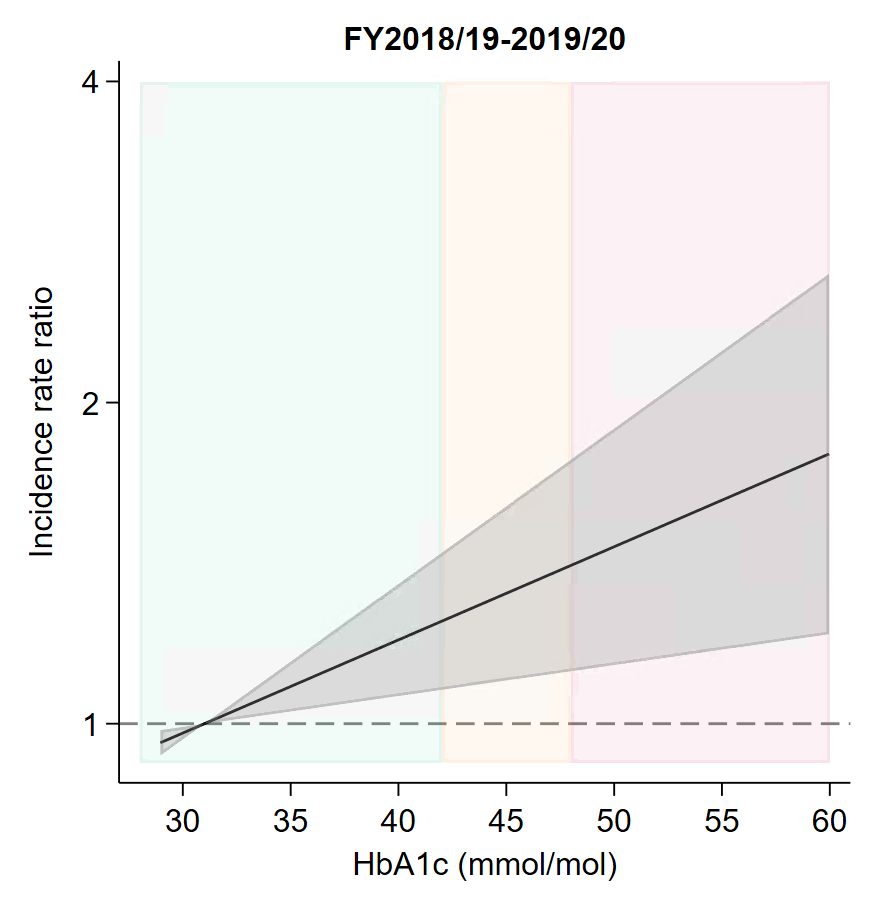

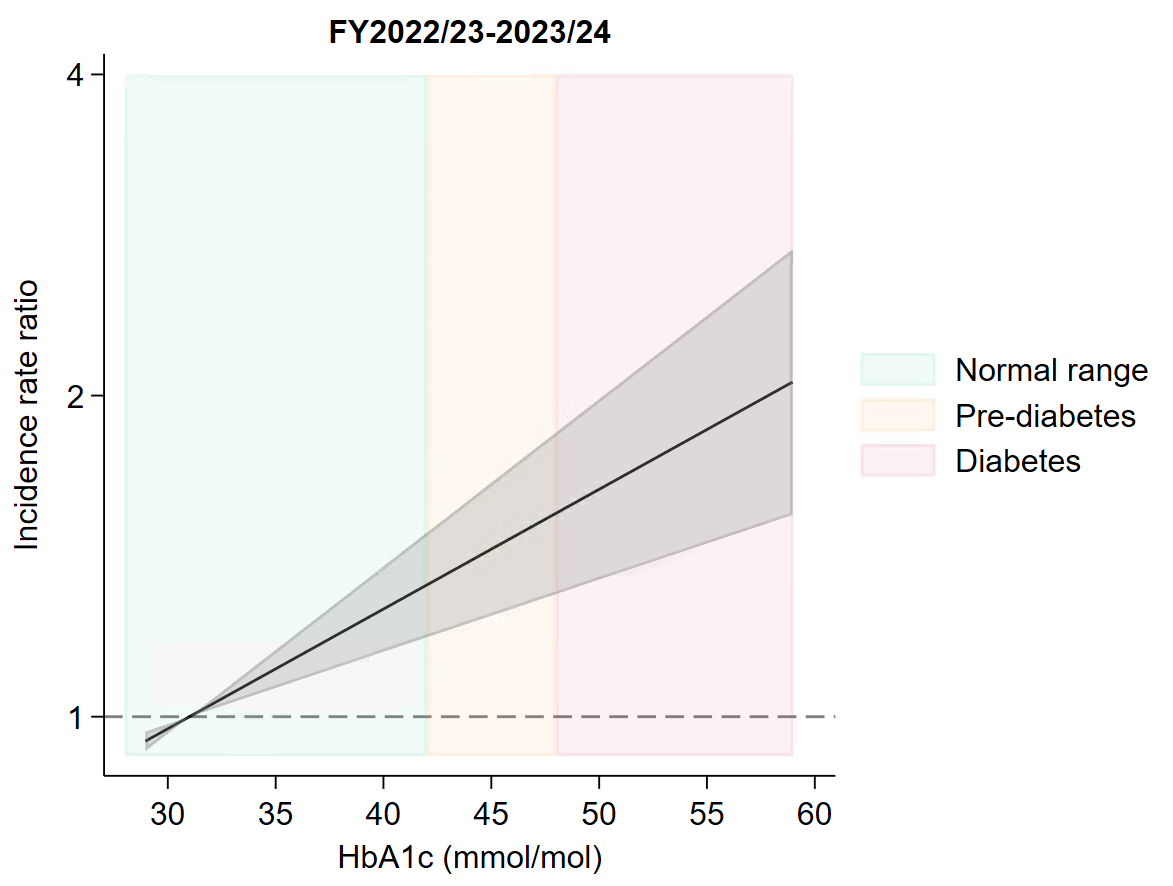

*Note: Normal range=below 42mmol/mol, prediabetes=42-47mmol/mol, diabetes, 48mmol/mol or over.^18^ FY2020/21-2021/22 not included as there was no evidence of an effect of HbA1c: IRR: 1·00 (95% CI: 0·99, 1·01; p-value = 0·731).*

Figure S7: Sensitivity and specificity of a hypothetical vaccine using different vaccination criteria in FY2018/19-2019/20, FY2020/21-2021/22, and FY2022/23-2023/24.

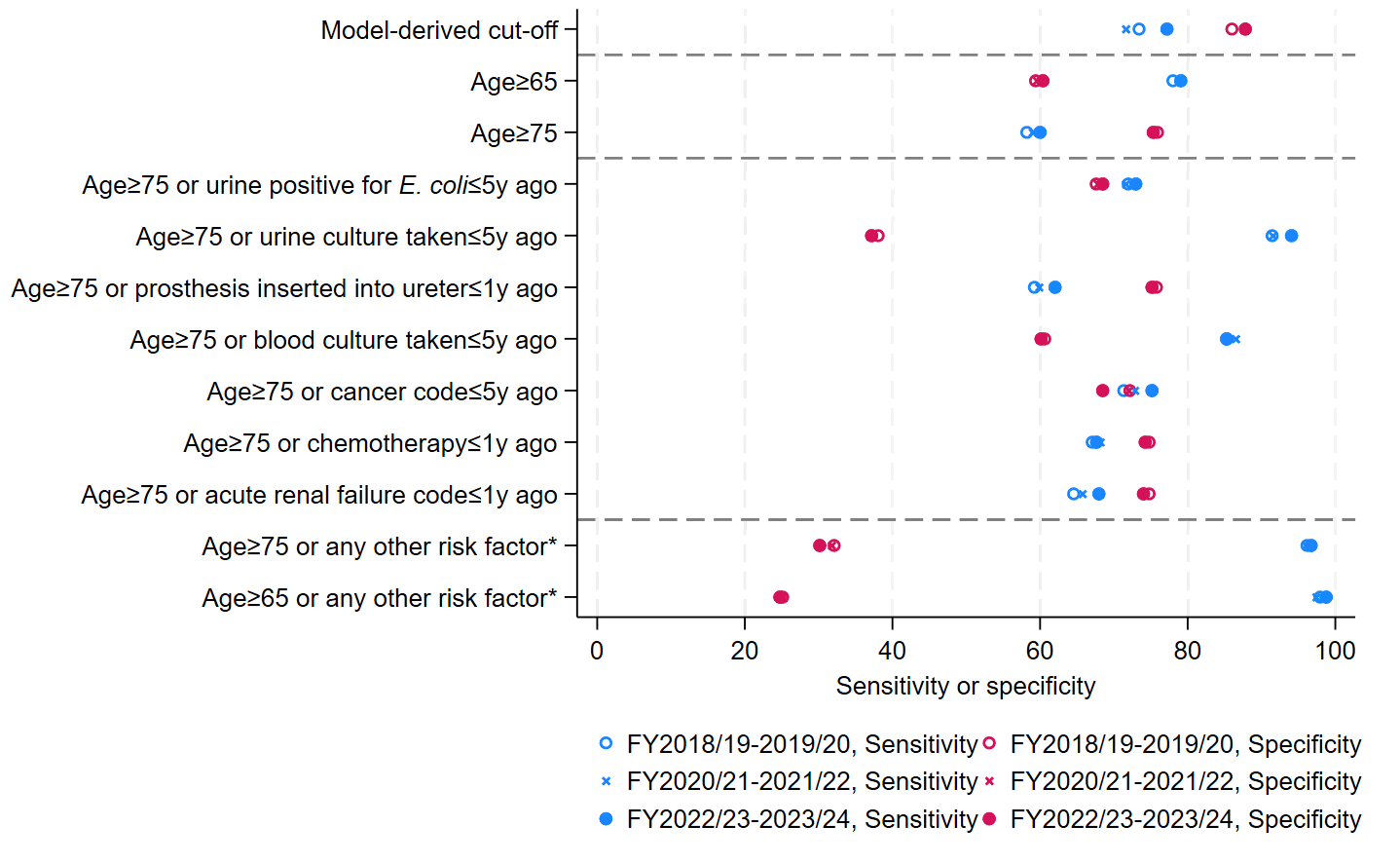

**Any of the seven individual risk factors presented in the main panel, specifically* *urine positive for E. coli (≤5y ago), urine culture taken (≤5y ago), prosthesis inserted into the ureter (≤1y ago), blood culture taken (≤5y ago), diagnosis code for cancer (≤5y ago), procedure code for chemotherapy (≤1y ago), or any diagnosis code for acute renal failure (≤1y ago).*

*Note: The FY2022/23-2023/24 variables and cut-off defined the “Model-derived cut-off” for all periods. For clarity, horizontal dashed lines separate model-derived cut-offs, age groups, specific risk factors, and all risk factors combined.*

Figure S8: Adjusted associations (incidence rate ratios with 95% CIs) between characteristics included as categorical or linear continuous variables and *E. coli* BSI risk in FY2020/21-2021/22.

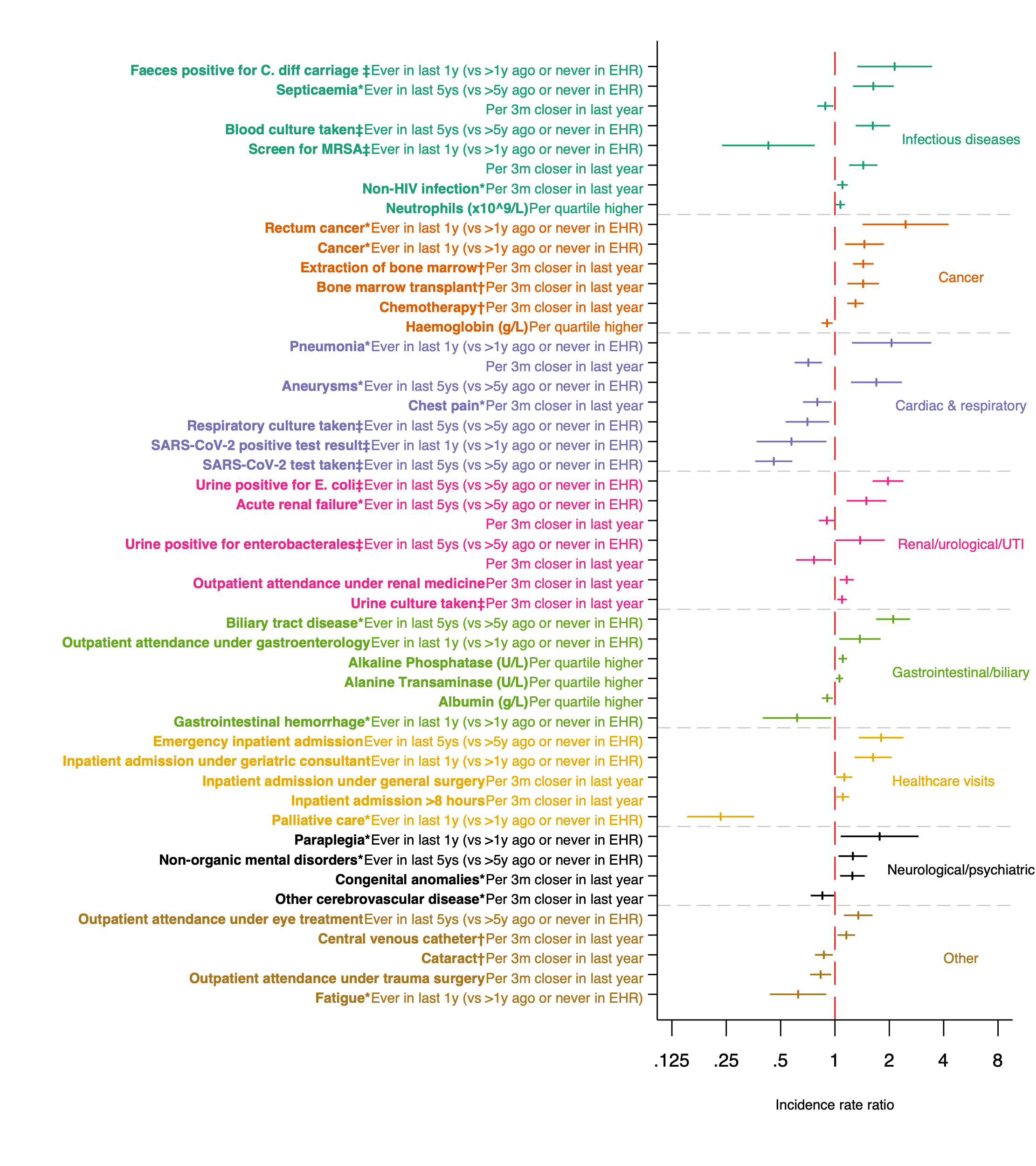
*Note: factor calculated from:* ********diagnosis codes,* ***†****procedure codes,* ***‡****microbiology data.*

Figure S9: Adjusted associations (incidence rate ratios with 95% CIs) between continuous characteristics with non-linear effects on *E. coli* BSI in FY2020/21-2021/22.

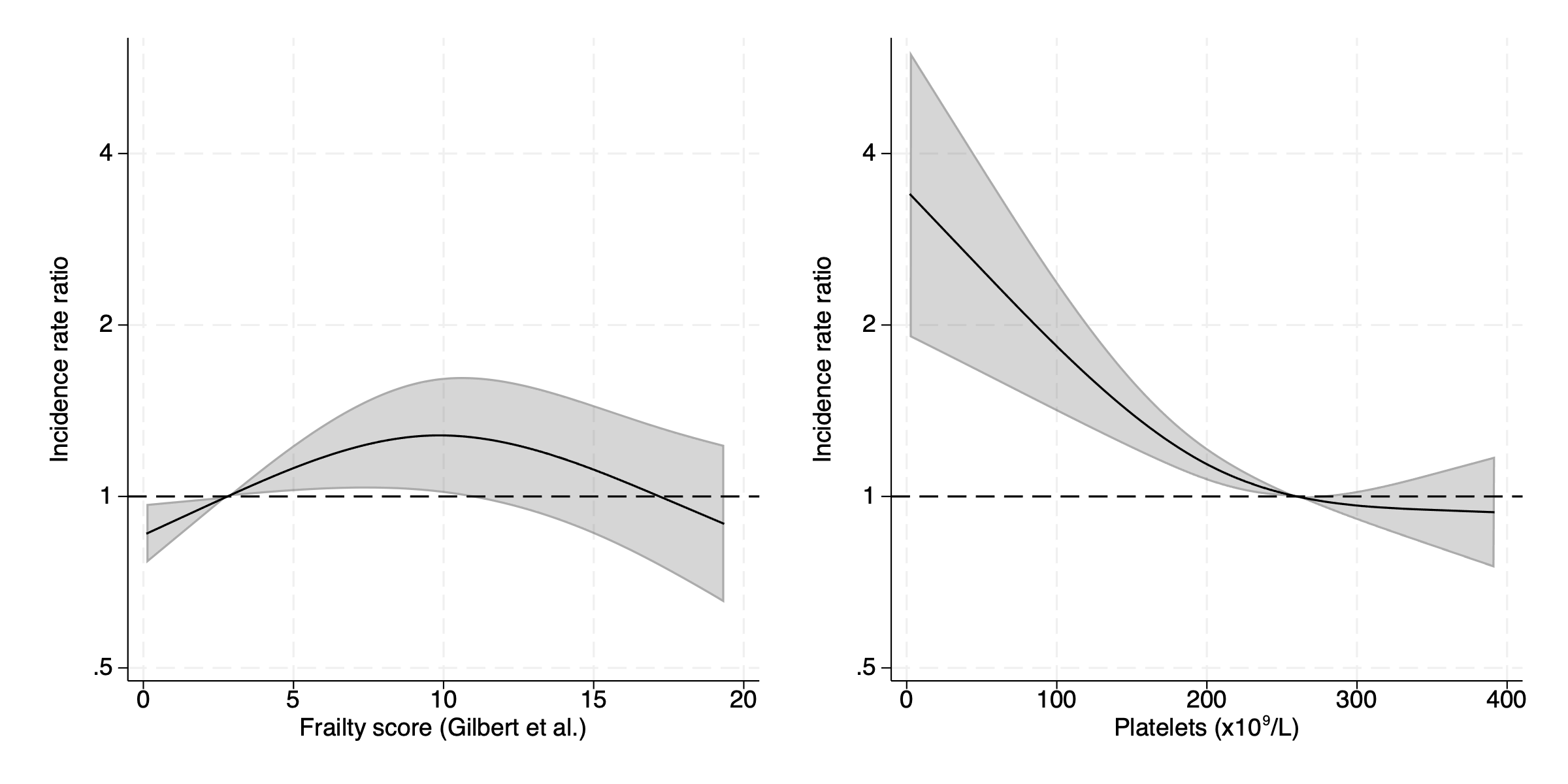

Figure S10: Incidence rate ratios with 95% CIs predicted from the final multivariate model from FY2020/21-FY2021/22 data for characteristics where a categorical variable and continuous linear variable were included.

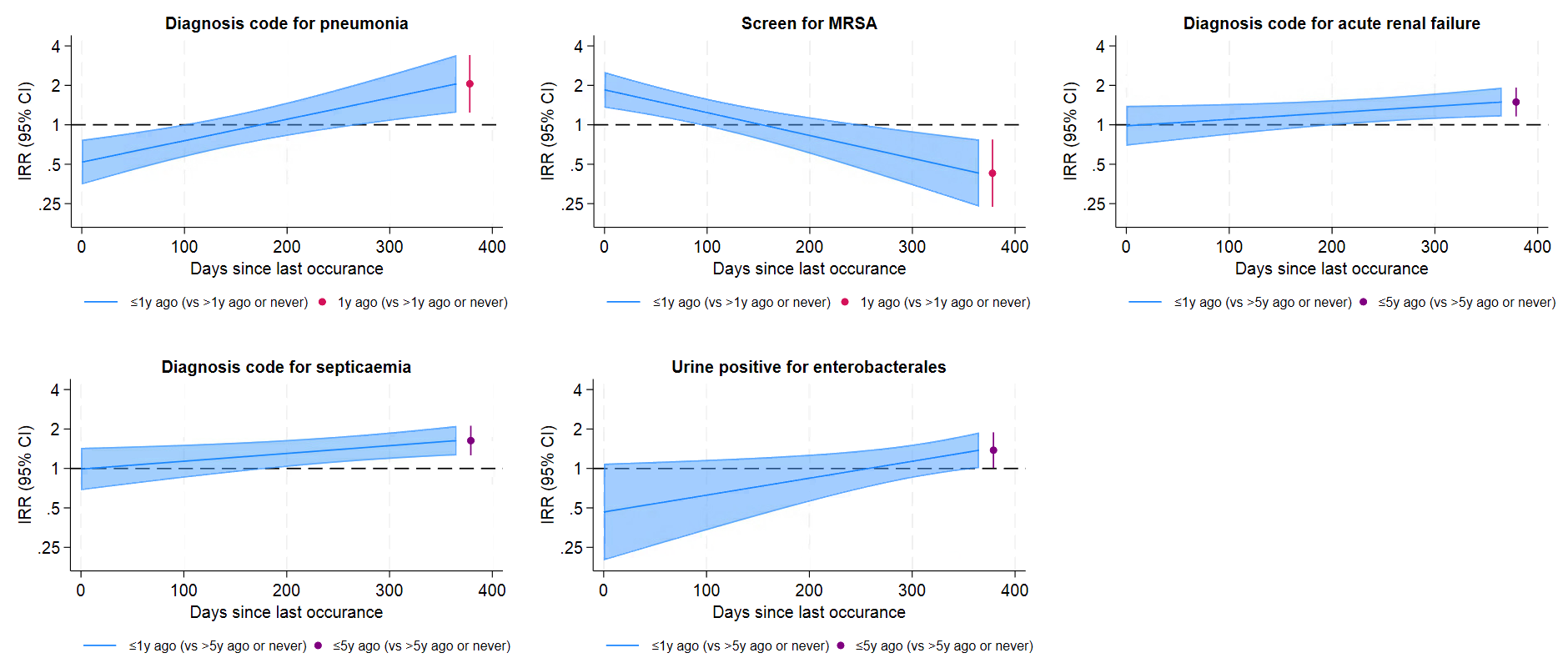

Figure S11: Adjusted associations (incidence rate ratios with 95% CIs) between characteristics included as categorical or linear continuous variables and *E. coli* BSI risk in FY2018/19-2019/20.

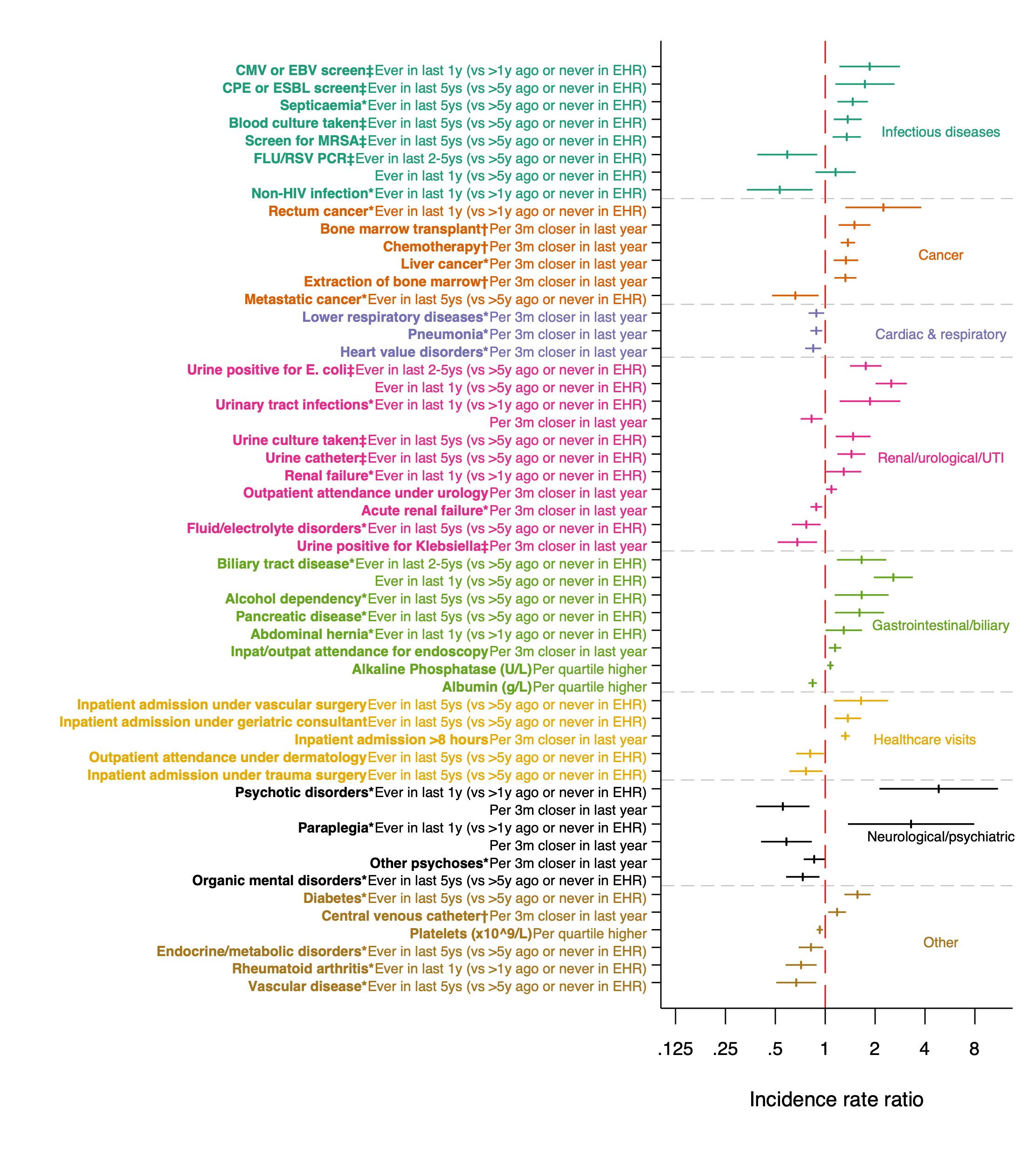

*Note: factor calculated from:* ********diagnosis codes,* ***†****procedure codes,* ***‡****microbiology data.*

Figure S12: Adjusted associations (incidence rate ratios with 95% CIs) between continuous characteristics with non-linear effects on *E. coli* BSI in FY2018/19-2019/20.

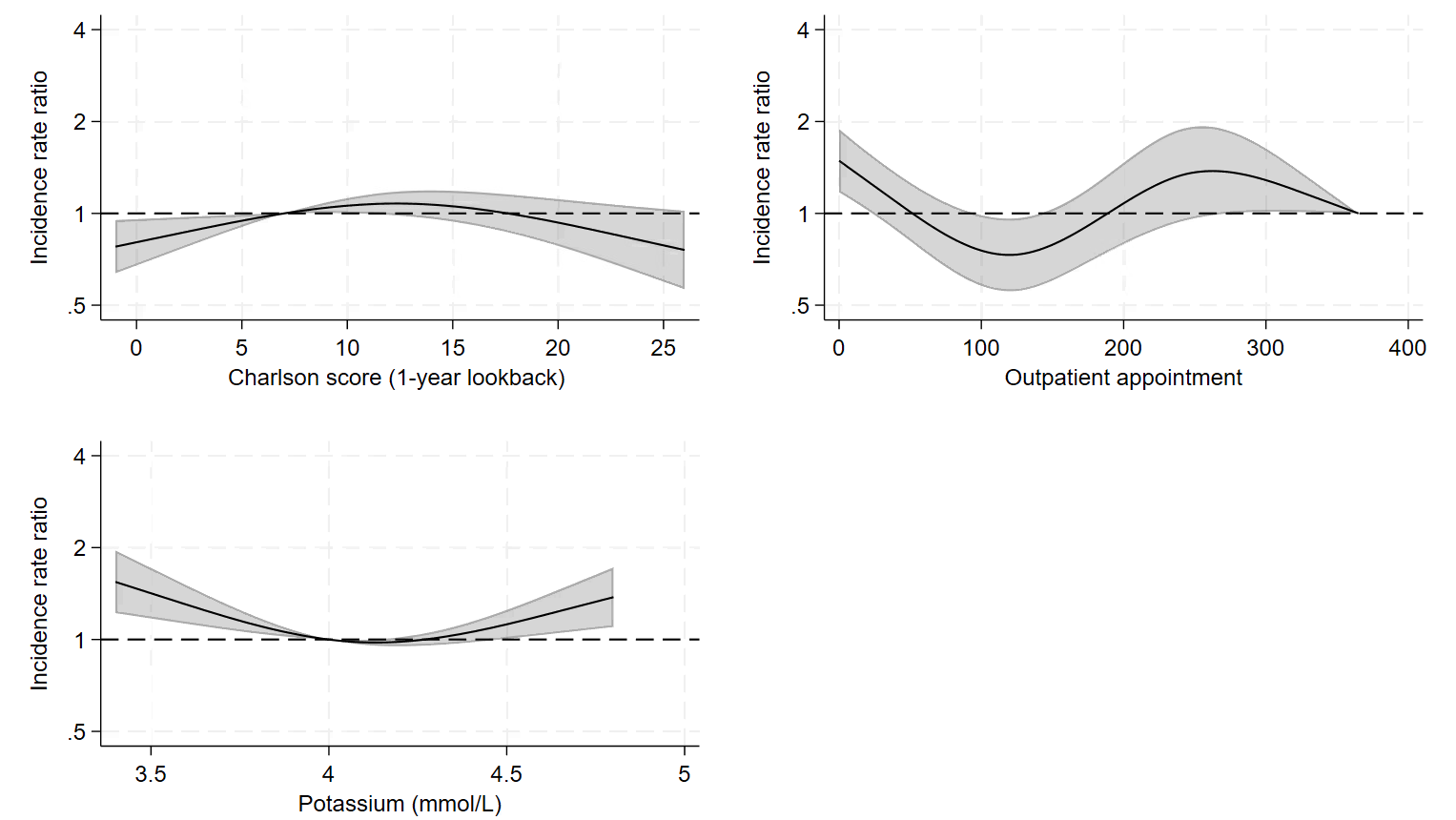

Figure S13: Incidence rate ratios with 95% CIs predicted from the final multivariate model from FY2018/19-2019/20 for characteristics where a categorical variable and continuous linear variable were included.

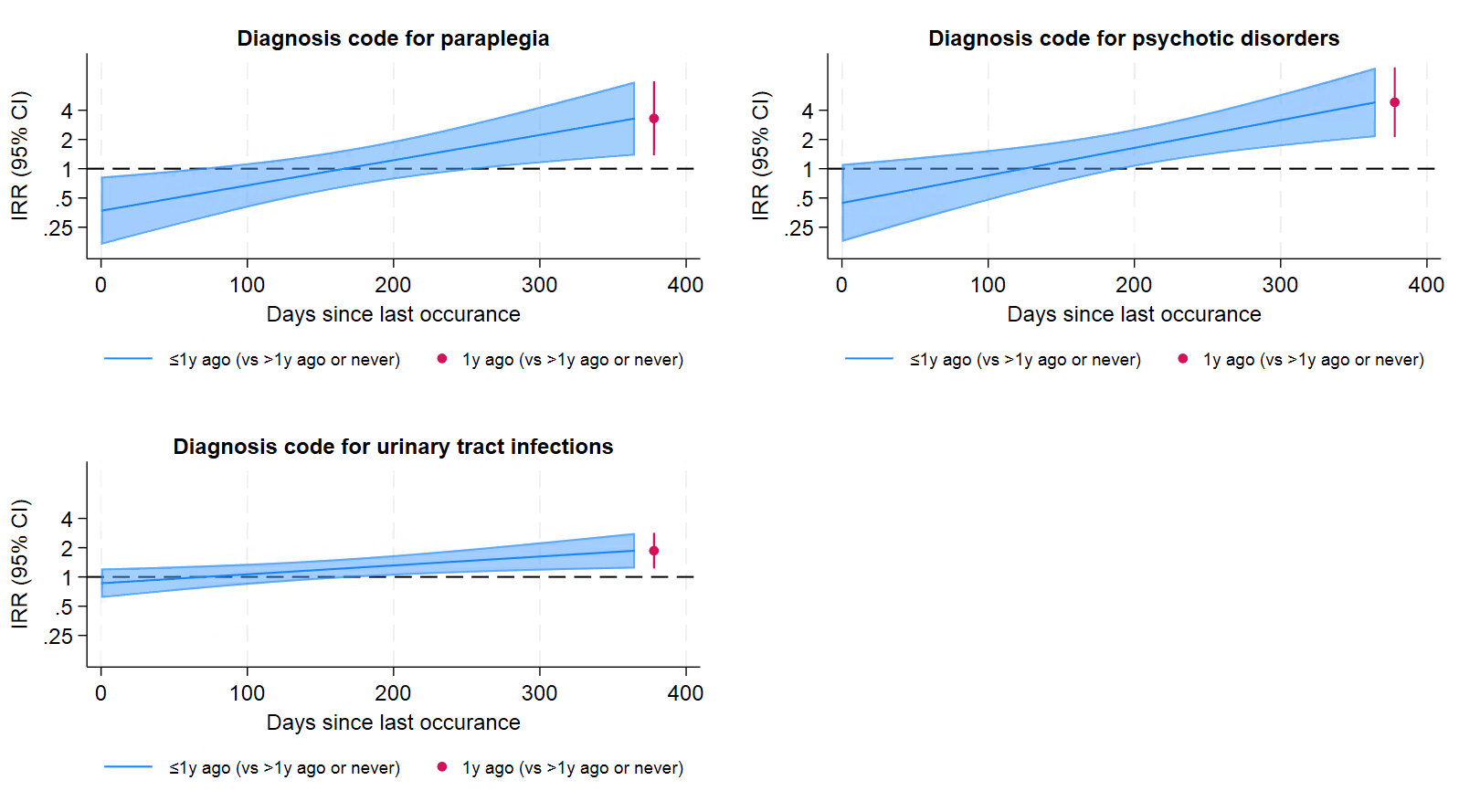

Figure S14: Prevalence of categorical factors selected for the multivariable model from FY2022/23-2023/24 across all three periods.

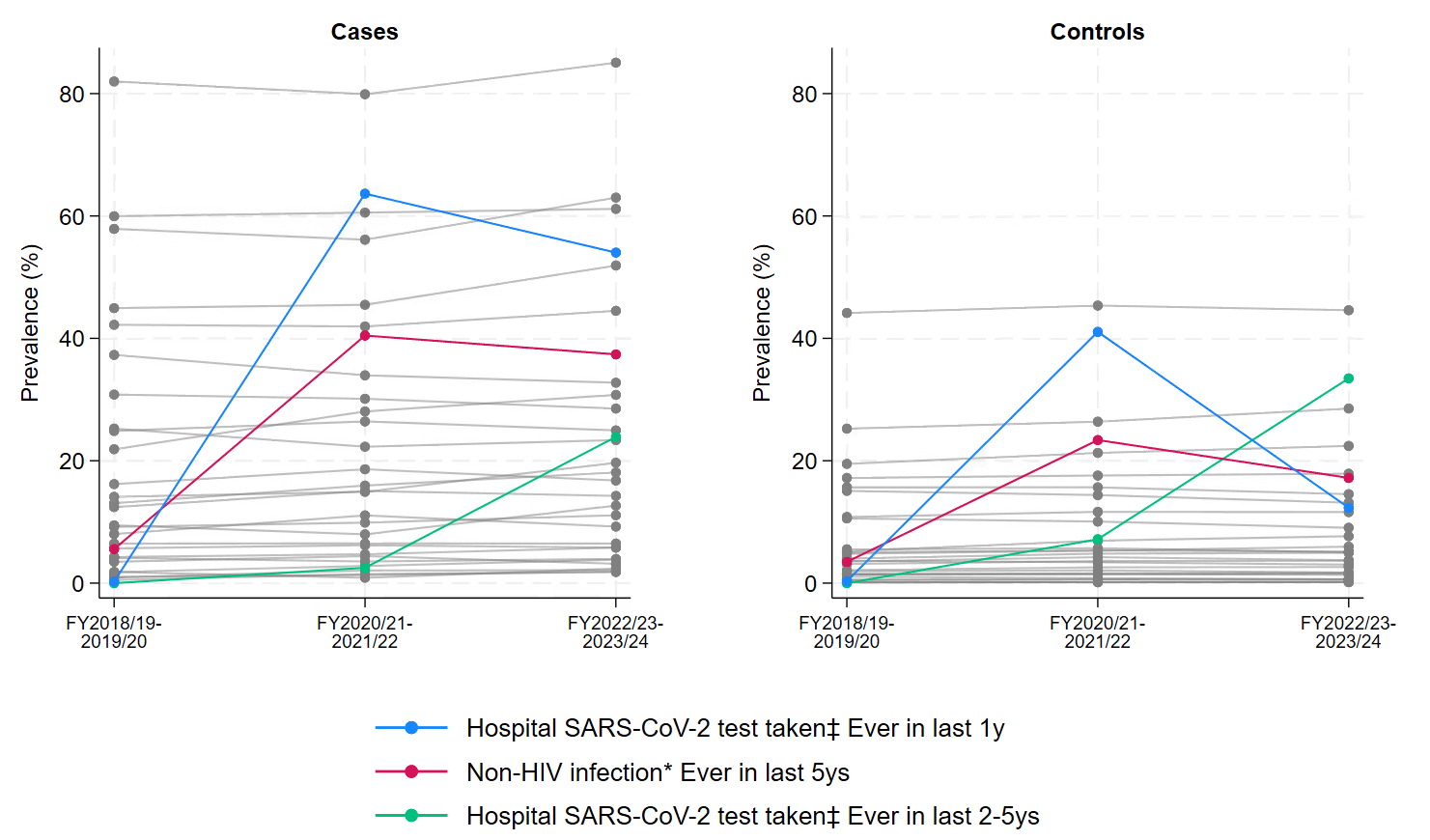

*Note: factor calculated from:* ********diagnosis codes,* ***†****procedure codes,* ***‡****microbiology data*

Figure S15: Median days since last occurrence for continuous factors selected for the multivariable model from FY2022/23-2023/24 across all three periods.

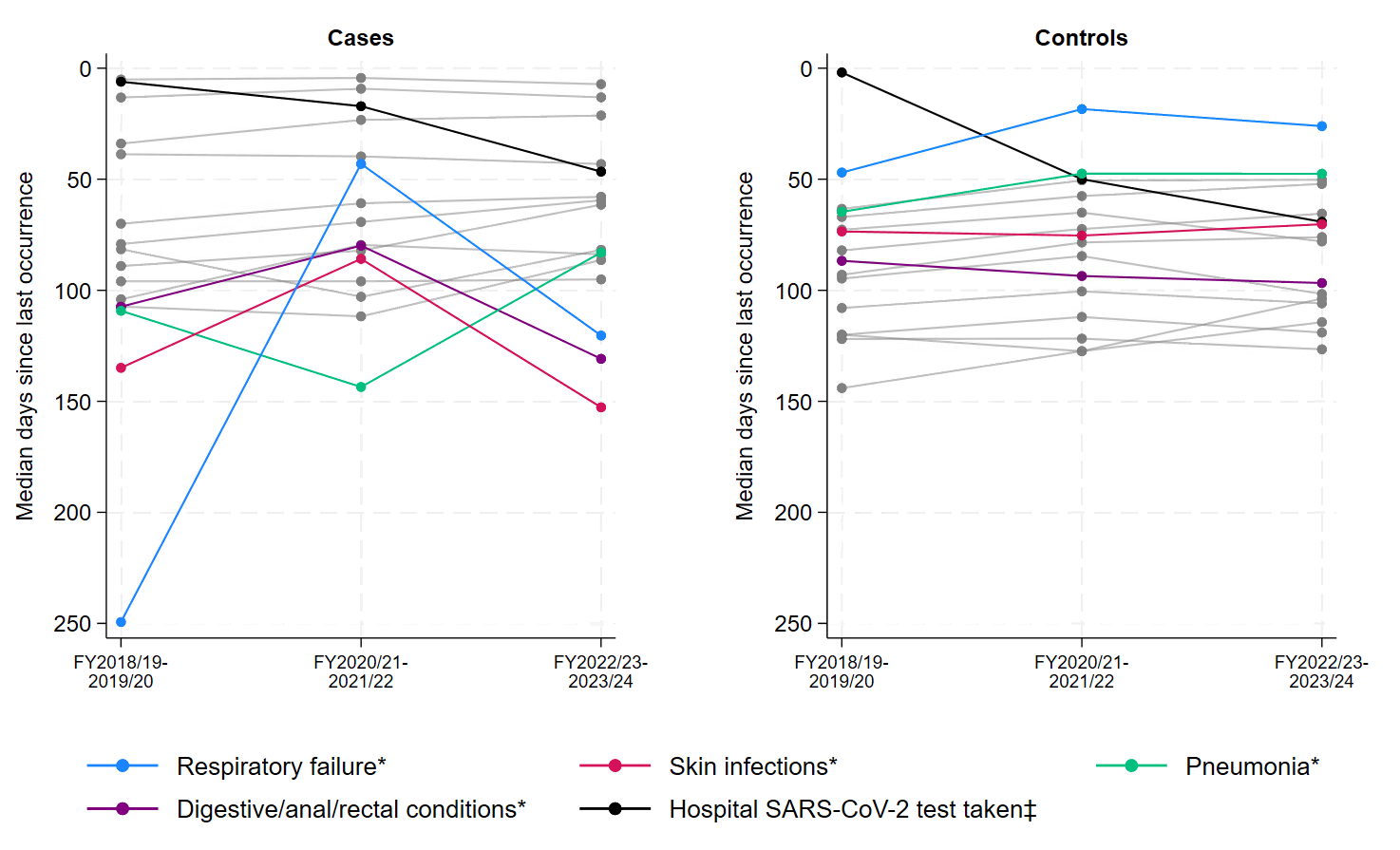

*Note: factor calculated from:* ********diagnosis codes,* ***†****procedure codes,* ***‡****microbiology data*

Figure S16: Median (IQR) for other continuous factors selected for the multivariable model from FY2022/23-2023/24 across all three periods.

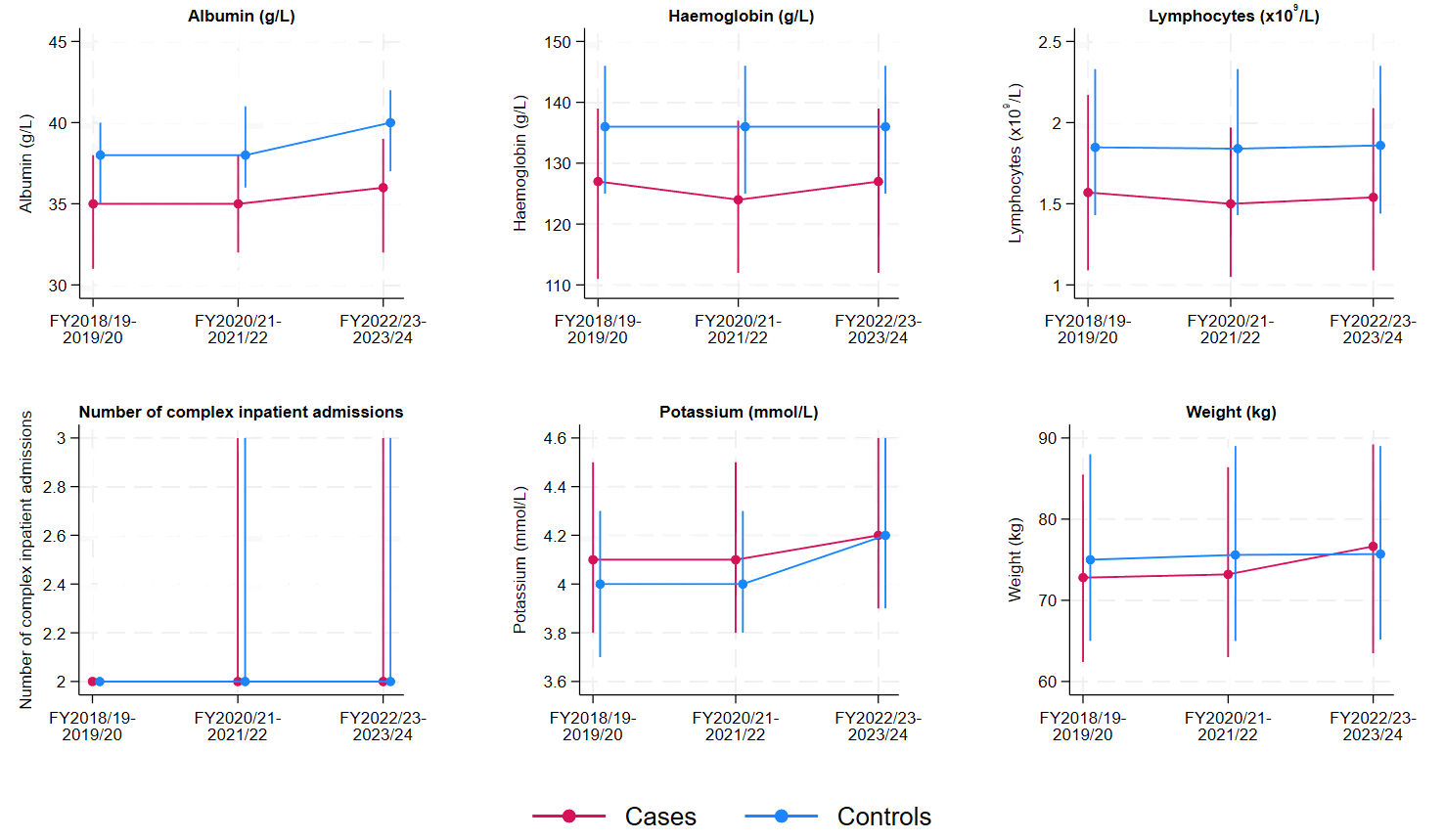

Figure S17: Percentage of cases prevented with varying vaccine effectiveness assumptions (60%, 75%, 90%).

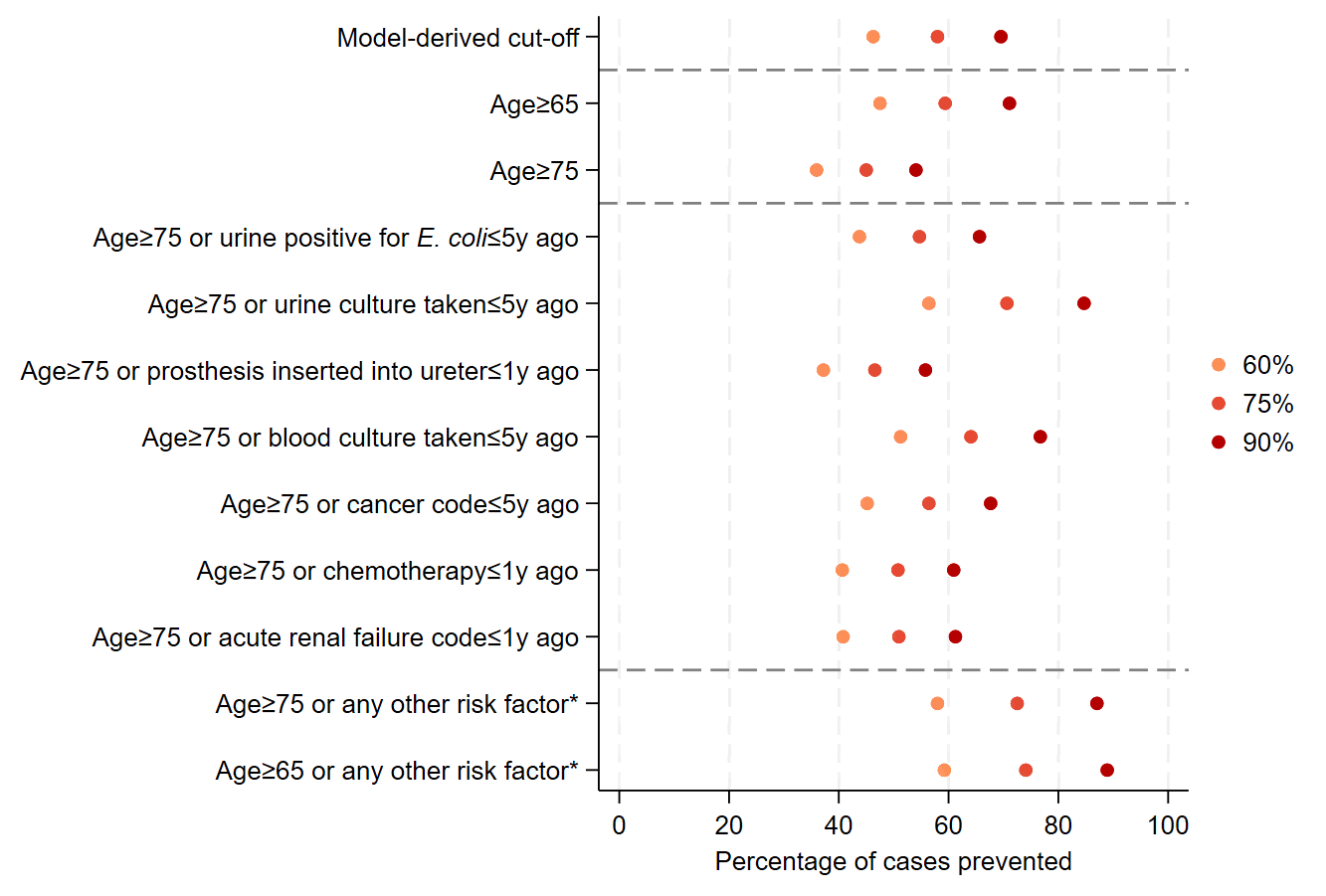

### 4 Supplementary Tables

Table S1: Distribution of core variables for cases (*E. coli* BSIs) and controls in FY2022/23-2023/24.

| **Characteristic** | **Cases, n (%) or median (IQR) [N=747]** | **Controls, n (%) or median (IQR) [276,758]** | **Cases removed from analysis (no prior inpatient episode in the last 5y) [N=196]** |
| --- | --- | --- | --- |
| **Age (years)** | 78 (67, 86) | 57 (38, 73) | 69 (54, 79) |
| Missing | 0 (0) | 0 (0) |  |
| **Sex** |  |  |  |
| Female | 361 (48) | 156,018 (56) | 105 (54) |
| Male | 396 (52) | 120,740 (44) | 91 (46) |
| Missing | 0 (0) | 0 (0) | 0 (0) |
| **Ethnicity** |  |  |  |
| White | 612 (81) | 190,546 (69) | 115 (59) |
| Non-white | 46 (6) | 21,489 (8) | 13 (7) |
| Missing | 99 (13) | 64,723 (23) | 68 (35) |
| **Deprivation percentile** | 74 (54, 89) | 74 (54, 89) | 70 (46, 85) |
| Missing | 4 (1) | 2,567 (1) | 13 (7) |
| **Rural/urban classification** |  |  |  |
| Urban city/town | 498 (66) | 181,153 (65) | 126 (64) |
| Town/fringe | 132 (17) | 44,813 (16) | 32 (16) |
| Rural village | 123 (16) | 48,219 (17) | 29 (15) |
| Missing | 4 (1) | 2,573 (1) | 9 (5) |
| **Catchment percent** | 95 (89, 96) | 93 (62, 96) | 88 (6, 95) |
| Missing | 4 (1) | 2,567 (1) | 13 (7) |

*Note: For deprivation percentile, 1=most deprived, 100=least deprived. Catchment percentile is the percentage of individuals in the local area visiting an Oxfordshire hospital as defined by the Office of Health Disparities; 0 = none, 100 = all.^2^*

Table S2: Variables with evidence of collinearity selected after backwards elimination and using p<0.25 as the entry threshold.

| **Characteristic** | **Level** | **Univariate IRR (95% CI) [p-value]** | **Univariate global p-value** | **Multivariate IRR (95% CI) [p-value]** | **Multivariate global p-value** |
| --- | --- | --- | --- | --- | --- |
| **Potentially related to collider bias / competing risks** | | | | | |
| Diagnosis code for fluid/electrolyte disorders (categorical) | In last 5Y (vs >5Y ago or never in EHR) | 2.39 (1.99, 2.87) | <0.001 | 0.73 (0.60, 0.89) | 0.02 |
| Diagnosis code for surgical wound risk | Per 3 months closer in last year | 1.23 (1.17, 1.30) | <0.001 | 0.91 (0.85, 0.97) | 0.003 |
| Emergency department visit | In last 1Y (vs >1Y ago or never in EHR) | 2.18 (1.86, 2.56) | <0.001 | 0.79 (0.65, 0.96) | 0.025 |
| **Potentially related to survivor bias** | | | | | |
| Number of complex inpatient admissions | Per 2 units higher | 4.61 (3.32, 6.40) | <0.001 | 0.52 (0.34, 0.80) | 0.003 |
| Complex inpatient admission | In last 5Y (vs >5Y ago or never in EHR) | 4.41 (3.18, 6.13) | <0.001 | 0.50 (0.32, 0.77) | 0.002 |
| **Potentially due to high correlation with another factor in the multivariate model** | | | | | |
| Diagnosis code for COVID-19 (categorical) | In last 5Y (vs >5Y ago or never in EHR) | 2.46 (1.95, 3.10) | <0.001 | 0.65 (0.46, 0.90) | 0.010 |

Table S3: Number of cases and controls included in the final multivariable model with and without HbA1c.

|  | **FY2018/19-2019/20** | | **FY2022/23-2023/24** | |
| --- | --- | --- | --- | --- |
| Individuals included | Controls, n (%) (N=221,610) | Cases, n (%) (N=687) | Controls, n (%) (N=210,401) | Cases, n (%) (N=654) |
| Final multivariable model | 190,167 (86) | 677 (99) | 167,410 (80) | 640 (98) |
| Final multivariable model plus HbA1c | 128,267 (58) | 544 (79) | 126,991 (60) | 565 (86) |

*Note: FY2020/21-2021/22 not included as there was no evidence of an effect of HbA1c: IRR: 1*·*00 (95% CI: 0*·*99, 1*·*01; p-value = 0*·*731).*

Table S4: BIC from models considering different combinations including a diagnosis code for COVID-19 and a SARS-CoV-2 positive test result from microbiology results.

| **Model specification** | **BIC** | **Model results** |
| --- | --- | --- |
| Both variables in the model (i.e. model after backwards elimination) | 7295.914 | Ever diagnosis code for COVID-19: IRR (95% CI) 0.65 (0.46, 0.90); p=0.010  Ever SARS-CoV-2 positive test result: IRR (95% CI) 1.48 (1.10, 2.00); p=0.010 |
| SARS-CoV-2 positive test result from microbiology data removed | 7289.015 | Ever diagnosis code for COVID-19: IRR (95% CI) 0.89 (0.70, 1.13); p=0.344 |
| Diagnosis code for COVID-19 removed | 7289.343 | Ever SARS-CoV-2 positive test result: IRR (95% CI) 1.09 (0.87, 1.36); p=0.443 |
| Combined variable which is any evidence of COVID-19/SARS-CoV-2 included | 7289.895 | Diagnosis code for COVID-19 or SARS-CoV-2 positive test: 1.00 (0.80, 1.24); p=0.927 |
| Both variables removed | 7277.871 | - |

*Note: All models included core variables and all screening variable selected after backwards elimination.*

Table S5: Summary of the number of variables selected after backward elimination across all FYs.

| Total number of periods present | Financial years present | n (%) (N=113) |
| --- | --- | --- |
| Three (N=11, 10%) | FY2018/19-2019/20, FY2020/21-2021/22, and FY2022/23-2023/24 | 11 (10) |
| Two (N=19, 17%) | FY2018/19-2019/20 and FY2020/21-2021/22 | 7 (6) |
|  | FY2018/19-2019/20 and FY2022/23-2023/24 | 6 (5) |
|  | FY2020/21-2021/22 and FY2022/23-2023/24 | 6 (5) |
| One (N=83, 73%) | FY2018/19-2019/20 | 29 (26) |
|  | FY2020/21-2021/22 | 26 (23) |
|  | FY2022/23-2023/24 | 28 (25) |

Table S6: Pseudo R-squared for models including variables identified or the final model in all three periods fit on all three data periods.

|  | **Pseudo R-squared (%)** | | |
| --- | --- | --- | --- |
| **Dataset** | FY2018/19-2019/20 variables | FY2020/21-2021/22 variables | FY2022/23-2023/24variables |
| FY2018/19-2019/20 | **22.1** | N/A | N/A |
| FY2020/21-2021/22 | 18.2 (-3.1) | **21.3** | 19.0 (-2.3) |
| FY2022/23-2023/24 | 19.5 (-3.6) | 19.2 (-3.9) | **23.1** |

References

1.	Ministry of Housing, Communities & Local Government,. The English Indices of Deprivation 2019 (IoD2019) 2019.

7.	Dr Foster Intelligence. Understanding HSMRs. A Toolkit on Hospital Standardised Mortality Ratios. 2012.

13.	UK Health Security Agency. Operating Procedure Codes Supplement (OPCS). Surgical Site Infection Surveillance Service (SSISS). 2019.

14.	Agniel D, Kohane IS, Weber GM. Biases in electronic health record data due to processes within the healthcare system: retrospective observational study. *BMJ* 2018; **361**: k1479.

15.	NHS inform. Common blood tests. 2023. <https://www.nhsinform.scot/tests-and-treatments/blood-tests/common-blood-tests/> (accessed 31 May 2024).

16.	Raftery A. Sociological methodology, chapter Bayesian Model Selection in Social Research Cambridge, MA: Wiley-Blackwell; 1998.

17.	Greenland S, Drescher K. Maximum likelihood estimation of the attributable fraction from logistic models. *Biometrics* 1993; **49**(3): 865-72.

18.	Diabetes.co.uk. Guide to HbA1c. 2023. <https://www.diabetes.co.uk/what-is-hba1c.html> (accessed 11 March 2025).
